# Cognitive Mechanisms of Aberrant Self-Referential Social Perception in Psychosis and Bipolar Disorder: Insights from Computational Modeling

**DOI:** 10.1101/2024.03.30.24304780

**Authors:** Carly A. Lasagna, Ivy F. Tso, Scott D. Blain, Timothy J. Pleskac

**Author notes:** Corresponding author. Address: Department of Psychology, University of Michigan, 530 Church St, Ann Arbor, MI 48109, USA. Denotes shared senior authorship (Ivy F. Tso, Timothy J. Pleskac).

## Abstract

**Background and Hypothesis:** Individuals with schizophrenia (SZ) and bipolar disorder (BD) show disruptions in self-referential gaze perception—a social perceptual process related to symptoms and functioning. However, our current mechanistic understanding of these dysfunctions and relationships is imprecise.

**Study Design:** The present study used mathematical modeling to uncover cognitive processes driving gaze perception abnormalities in SZ and BD, and how they relate to cognition, symptoms, and social functioning. We modeled the behavior of 28 SZ, 38 BD, and 34 controls (HC) in a self-referential gaze perception task using drift-diffusion models (DDM) parameterized to index key cognitive components: drift rate (evidence accumulation efficiency), drift bias (perceptual bias), start point (expectation bias), threshold separation (response caution), and non-decision time (encoding/motor processes).

**Study Results:** Results revealed that aberrant gaze perception in SZ and BD was driven by less efficient evidence accumulation, perceptual biases predisposing self-referential responses, and greater caution (SZ only). Across SZ and HC, poorer social functioning was related to greater expectation biases. Within SZ, perceptual and expectancy biases were associated with hallucination and delusion severity, respectively.

**Conclusions:** These findings indicate that diminished evidence accumulation and perceptual biases may underlie altered gaze perception in patients and that SZ may engage in compensatory cautiousness, sacrificing response speed to preserve accuracy. Moreover, biases at the belief and perceptual levels may relate to symptoms and functioning. Computational modeling can, therefore, be used to achieve a more nuanced, cognitive process-level understanding of the mechanisms of social cognitive difficulties, including gaze perception, in individuals with SZ and BD.

## Introduction

Individuals with schizophrenia (SZ) and bipolar disorder (BD) display chronic, medication-resistant functional impairments in social, familial, and role domains ^1–5^. Major drivers of functional impairments include persistent deficits in social cognition, which allow individuals to understand information about others ^6–8^ and occur regardless of illness phase ^9,10^. One critical determinant of social cognition is *self-referential gaze perception*—the ability to judge whether others are looking at us ^11,12^. Gaze information is used with facial emotion and head orientation cues to understand others’ inner states ^13^, informing decision-making, behavior, and functioning ^14,15^. Yet, in SZ and BD, these abilities are disrupted. Patients make slower ^16–21^, less accurate ^17,20,22^, and/or less precise judgments about others’ gaze ^23–27^, and show self-referential biases toward over-endorsing eye contact ^23–26,28,29,29,30^. These relate to poorer general/social cognition ^16,23,23,24,27–29,29^ (in SZ and BD) and greater positive/negative symptoms (in SZ)^22,23,26,28^.

The clinical and functional relevance of gaze perception deficits in SZ and BD demand a deeper mechanistic account. Yet, perceptual choices like gaze perception depend on several intertwined processes—including expectations, encoding, evidence accumulation, response caution, and motor execution ^31^—that traditional accuracy and reaction time (RT) measures cannot disentangle. One reason for this, as detailed below, is that multiple explanatory pathways can account for a single disruption in gaze perception. This is important for SZ and BD, which show overlapping (e.g., general cognitive deficits ^32^) *and* unique phenomenology (e.g., trait impulsivity in BD ^33,34^; mood-independent paranoia in SZ) that may predispose gaze perception disruptions through shared *and* distinct pathways. For instance, slower and less accurate choices may arise from (cognition-related) inefficient evidence accumulation in both groups, (impulsivity-related) reduced caution in BD, and/or (paranoia-related) expectancy biases in SZ.

Computational modeling provides a solution to characterizing these different processes. We employed drift-diffusion models (DDM) to characterize the mechanisms of gaze perception disruptions in SZ and BD, and their relationships with general/social cognition, symptoms, and social functioning. According to the DDM, when deciding whether someone is looking at them or not, individuals accumulate noisy sensory evidence about both choices (Figure 1A). A decision is made once evidence reaches a threshold for either choice.

**Figure 1.**
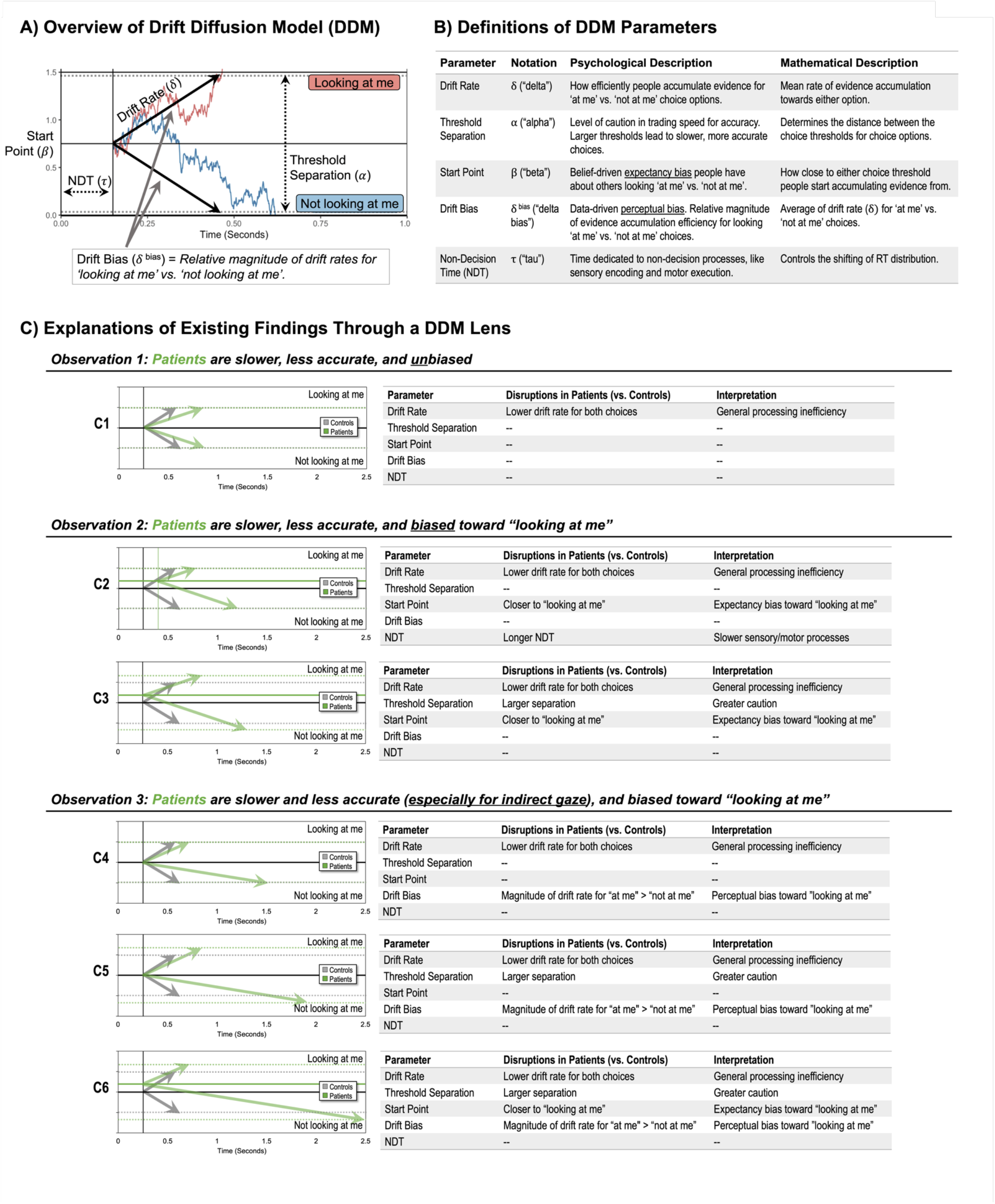
Overview of DDM and DDM accounts of extant findings. DDM application to gaze perception; B) Definitions of DDM parameters; C) Several plausible hypotheses that could account for existing data on gaze perception in SZ and BD. Dashed horizontal lines designate decision boundaries, where the vertical distance between decision boundaries represents the threshold separation (i.e., response caution) parameter.

Through an extensive history of testing and experimental validation ^31,35,36^, different cognitive processes have been linked to different DDM parameters (Figure 1B), supporting various pathways to perceptual disruptions (Figure 1C). The *drift rate* determines the direction and rate of evidence accumulation, indexing how efficiently evidence is accumulated to determine if someone is looking at them. Lower drift rates mean slower, more error-prone choices (Figure 1.C1-C6). The separation between choice thresholds determines how much evidence is needed before deciding. Higher separation means more evidence and, consequently, slower and less error-prone choices (Figure 1.C3/C5-C6). Thus, *threshold separation* determines cautiousness in trading speed for accuracy. Two DDM mechanisms can produce self-referential biases. First, if the *start point* for evidence accumulation begins closer to one choice threshold, responses will be biased toward and faster for that choice (Figure 1.C3/C6). Thus, it determines initial expectancy biases for detecting self-referential gaze. Second, is the *drift bias*: a relative asymmetry in drift rate magnitudes for direct and indirect gaze. If participants process “looking at me” information more efficiently, this can reduce the evidence accumulation rate for indirect gaze, creating a perceptual bias toward self-referential responses (Figure 1.C4-C6). Finally, *non-decision time* (NDT) captures sensory and motor processes, separate from the deliberation process. Longer NDT’s mean longer RTs (Figure 1.C2).

We leveraged extant data ^16,17,20,21^ to characterize cognitive processes driving gaze perception using a DDM framework ^37^. We hypothesized that SZ and BD would show less efficient evidence accumulation than HC because patients exhibit lower drift rates across cognitive tasks ^38–44^. We explored group differences in other parameters to delineate among mechanistic explanations in Figure 1C. Finally, we examined the mechanisms of relationships between gaze perception and general/social cognition, symptoms, and social functioning by exploring associations with DDM parameters.

## Methods

### Participants

The present study re-examined two existing datasets (used in prior publications ^16,17,20,21^) consisting of 28 SZ, 38 BD, and 34 HC participating in two waves of a larger study examining social processing in SZ (“SZ sub-study” [28 SZ, 18 HC]) and BD (“BD sub-study” [38 BD, 16 HC]). Details about recruitment, inclusion/exclusion criteria, and data collection are in Supplement 2-3.

### Measures

#### Assessments

Diagnoses were confirmed using the Structured Clinical Interview for DSM-IV-TR ^45^ or Diagnostic Interview for Genetic Studies ^46^. In the SZ sub-study, SZ symptoms (Scale for Assessment of Positive Symptoms [SAPS ^47^]; Scale for Assessment of Negative Symptoms [SANS ^48^]), depressive symptoms (Beck Depression Inventory-II [BDI-II ^49^]), general cognition (Brief Assessment of Cognition in Schizophrenia [BACS] ^50,51^), emotion-related social cognition (Mayer-Salovey-Caruso Emotional Intelligence Test [MSCEIT] ^52^), and social functioning (Social Adjustment Scale Self-Report, Social/Leisure scale [SASSR-Social] ^53,54^; inverse-coded) were assessed. In the BD sub-study, mania (Altman Self-Rated Mania scale [ASRM ^55^]) and depressive symptoms (BDI-II) were assessed. Details about inter-rater reliability and assessment scoring are in Supplement 4.

We focused on three SZ symptom domains—delusions, hallucinations, and negative symptom-related amotivation. Hallucinations may stem from initial biases applied to perceptual experiences ^56^, while delusions may stem from biases at the belief level ^57,58^. Both can potentially influence gaze perception. Because DDM parameters can capture belief and perceptual biases through the start point and drift bias ^59^, respectively, we examined relationships between SAPS- Hallucinations, SAPS-Delusions, and DDM parameters to examine the potential mechanisms of relationships between gaze perception and positive symptoms in SZ. Evidence supports a 2- factor structure of negative symptoms consisting of amotivation (avolition, anhedonia, asociality) and diminished expression (flat affect, alogia) ^60–64^. Because the former relates to social cognition, general functioning ^62^, and task performance ^65–67^, we also examined relationships between a negative symptom amotivation factor ^68^ and DDM parameters.

Recent work highlights links between social cognition with depressive and hypo/manic symptoms ^6,10,69^, but links between mood symptoms and gaze perception *specifically* are less well-understood in SZ and/or BD. Thus, we explored relationships between DDM parameters and ASRM-Mania and BDI-Depression.

#### Gaze perception task

Participants completed a self-referential gaze perception task ^16^ in which they indicated whether faces were looking at them (Yes) or not (No) via button press (Figure 2). Stimuli were greyscale face images ^70^ depicting different gaze directions (direct, indirect), emotions (neutral, fearful), and head orientations (forward, deviated). Trials consisted of a 1000ms fixation, 100ms blank, 100ms stimulus, up to 2000ms response period (terminated following response), and a 600ms inter-trial interval. Trials were presented randomly across four blocks (64 trials per condition x 8 conditions = 512 trials). Block order (neutral-fearful-neutral-fearful -or- fearful-neutral-fearful-neutral) and response orientation (yes-CTRL, no-SHIFT -or- yes-SHIFT, no- CTRL) were counterbalanced across participants. Participants began with a brief practice to acclimate to the task. The task was programmed in E-prime 2.0 Professional and lasted ∼20-25 minutes. Non-response trials and trials with invalidly quick responses (< 250ms) ^71^ were removed.

**Figure 2.** Gaze discrimination task. Participants pressed a button to indicate whether the face was looking at them (Yes/No). Faces varied in gaze direction (direct, averted), emotion (neutral, fearful), and head orientation (forward, deviated). ITI = inter-trial interval; ms = milliseconds.

To index performance, in addition to deriving DDM parameters, we calculated two sets of traditional behavioral measures. First, we calculated task-averaged reaction time (RT) and accuracy. Second, although the DDM is considered a dynamic extension of traditional signal detection theory (SDT) ^72^—that can identify multiple sources of bias by modeling choices *and* RTs—traditional SDT is more common in psychopathology research. To facilitate comparisons with previous research, we used an equal variance Gaussian SDT model in Stan (see Supplement 16) to estimate two traditional SDT measures: SDT-Discriminability (sensitivity in discriminating between choices) and SDT-Criterion (threshold for decision criteria).

### Computational modeling

We used hierarchical Bayesian DDM’s ^37^ to characterize the mechanisms of gaze perception in our three groups, using an exploratory modeling-building approach to determine a model that best accounted for choices and RTs. We parameterized the DDM to examine how diagnostic group, others’ gaze direction, head orientation, and emotional expression impact the computationally defined processes laid out by the DDM during gaze perception. In the following sections, each modeling step is summarized and the reader is directed to specific supplement sections containing complete descriptions of these procedures and results.

#### 1) Model specification

We began with a full model space of 30+ variations of the DDM. Critically, these differed in their specification of how task conditions (gaze, head orientation, and emotion of stimuli) impacted DDM parameters. This model space was then narrowed to *n* = 8 plausible models (see Supplement 5), which were tested according to recommended guidelines ^73^. All were hierarchical Bayesian models where subject-level fixed effects for all parameters were informed by (diagnostic) group-level means and variances. We used weakly informative priors with a non-centered parameterization (i.e., parameters are sampled in a ‘standard’ space and later scaled/transformed), which can help sampling for complex models (see Supplement 6 for model specifications).

#### 2) Model implementation

Models were implemented in Stan 2.21.0 ^74^ on a high-performance computing cluster using RStan 2.21.7 and cmdstanr 0.0.6 (cmdstan 2.32.2). We performed hierarchical Bayesian sampling using Stan’s No-U-Turn sampler, an adaptive variant of Hamiltonian Monte Carlo. Model testing and comparison were based on 8,000 post-warmup samples. After selecting a winning model, a final version was run using 36 chains with 2,500 warm-up samples and 6,000 post-warmup draws per chain, resulting in 216,000 total post-warmup draws.

#### 3) Convergence

Convergence checks performed for all models (see Supplement 7 and 13.1) suggested that parameters converged to target distributions. There were no divergences and, for all parameters, R-hat values were < 1.1 ^75^, trace plots were well-mixed, and autocorrelation was ∼0 by a lag of ∼30.

#### 4) Parameter recovery

Parameter recovery was performed on the baseline model and the winning model (see Supplement 8). Results showed good recovery of parameters for the baseline model (group-level: 94-98% recovery; subject-level: 94-96% recovery) and the winning model (group-level: 88- 100% recovery; 92-96% recovery).

#### 5) Model comparison

To determine which model best accounted for the data, we performed model comparisons using leave-one-out (LOO) cross-validation ^76^ (see Supplement 9). Cross-validation was used to avoid over-fitting ^73^. Differences in out-of-sample predictive accuracy were assessed based on changes in the expected log pointwise predictive density (ΔELPD-LOO; via ‘loo_compare’ in ^77^), where higher ELPD indicates better model fit. Results showed that the winning model (“Model 10” hereafter) for all groups was one in which all parameters varied by diagnostic group and evidence accumulation (drift rate) was influenced by gaze direction, head orientation, and emotion expression of stimuli. It assumed that response caution (threshold separation), start point (expectancy bias), and NDT operated as trait-level processes that did not vary in response to stimulus changes. This is a reasonable model account because the drift rate is influenced by the physical qualities of the stimulus.

#### 6) Confusion matrix

A confusion matrix helped assess whether we could arbitrate between models— particularly the winning model versus others (see Supplement 10). This evaluates whether data simulated from one model is best fit by the same model. Results showed it was possible to arbitrate between the winning model and others in 98% of simulations.

#### 7) Predictive checks

Posterior predictive checks tested the accuracy of predicted choices and RT distributions produced by the winning model (see Supplement 11). Results showed that predicted choice proportions and RT distributions mapped closely onto observed choice proportions and RT distributions for all diagnostic groups and task conditions. The exception is that the model slightly underpredicted the RT distribution tails when few trials were available, specifically when predicting RTs of incorrect responses in high-accuracy conditions. Together, this indicated that the winning model had high predictive accuracy.

#### 8) Additional data preparation

A preliminary examination of condition effects on DDM group-level parameters indicated that the congruency of head orientation with gaze direction credibly influenced drift rates, but facial emotion did not (Supplement 14.5-14.7). Therefore, for analysis of group differences (see ‘Statistical Analyses’ below), we marginalized drift rates for emotion conditions, meaning we averaged over each posterior sample for fearful and neutral faces within direct- forward, direct-deviated, indirect-forward, and indirect-deviated conditions. Then, for correlations and regressions, to make the number of tests more manageable, we calculated two measures—overall drift rates and drift biases—separately for forward and deviated heads.

Marginalizing over emotion conditions may appear counterintuitive when the winning model— accounting for the influences of gaze, head, *and* emotion on drift rates—outperformed other models that did not also account for the influence of emotion on drift rates. The winning model likely captured subtleties in how emotion influenced evidence accumulation that helped improve out-of-sample predictions but were not sufficiently large to yield credible condition-level effects.

Overall drift rates were calculated by flipping the sign of indirect-forward and indirect- deviated samples (originally negative-going) and averaging over each posterior sample for direct and indirect faces within forward and deviated head conditions. Drift bias was calculated using the same process, but *without* flipping the sign for indirect gaze posterior samples. This indexed the relative magnitude of drift rates for direct and indirect gaze, where drift bias values > 0 indicate greater relative efficiency for direct gaze (perceptual bias toward “looking at me”); and values < 0 indicate greater relative efficiency for indirect gaze (perceptual bias toward “not looking at me”).

### Statistical analyses

Using Bayesian statistics, analyses were performed on parameters from the winning model—Model 10—in R 4.1.0 (via RStudio 1.4.1717) ^78^. Before running analyses, to retain as much data as possible, we winsorized the outermost 1% of observations for measures that contained outliers. Because we used Bayesian tests based on posteriors, which do not change with the number of tests ^79,80^, there were no corrections for multiple comparisons.

#### Analysis plan

*First*, we examined between-group and within-group condition differences in DDM parameters using 90% highest density intervals (HDI) of posterior differences between groups and task conditions. We considered a 90% HDI of differences that did *not* contain zero a credible difference. We report 90% HDIs in brackets alongside the means of the difference interval.

*Second*, we examined potential mechanisms of relationships between gaze perception and general and social cognition found in prior studies. We ran exploratory Bayesian correlations (‘correlationBF’ in ^81^) between DDM parameters, BACS, and MSCEIT in the sub-sample for which BACS (28 SZ, 18 HC) and MSCEIT were collected (27 SZ, 18 HC). We did the same for relationships between DDM parameters and delusions (SAPS-Delusion; 28 SZ), hallucinations (SAPS-Hallucination), negative symptom-related amotivation (SANS-Amotivation; 27 SZ), depressive symptoms (BDI; 27 SZ, 36 BD, 27 HC), and mania symptoms (ASRM; 36 BD, 14 HC) using data from participants with complete data. We included traditional performance metrics in correlations (accuracy, RT, SDT-Criterion, SDT-Discriminability) to ascertain whether observed relationships were unique to DDM parameters. Additionally, because antipsychotics influence RTs, we tested whether antipsychotic dose (i.e., chlorpromazine equivalency [CPZeq]; calculated with ‘chlorpromazineR’ ^82^) correlated with performance- derived measures. We considered associations credible if the 90% HDI of the correlation coefficient did *not* contain zero. The strength of associations was interrogated with Bayes Factors (BF), which index the relative strength of evidence for the alternative (association exists) over the null (no association). Conventional BF interpretation ranges were used ^83^: *BF < 1 favors null* = 0.33–1 (anecdotal), 0.10–0.33 (moderate), 0.033–0.10 (strong), 0.01–0.033 (very strong), <0.01 (extreme); *BF > 1 favors alternative* = 1–3 (anecdotal), 3–10 (moderate), 10–30 (strong), 30–100 (very strong), >100 (extreme). We report posterior means and 90% HDIs of each r correlation coefficient, alongside the BF (i.e., “r = M, [HDI], BF = X”).

*Third,* to maximize our sample size we combined data of 27 SZ and 18 HC with complete SASSR-Social data and tested whether DDM parameters could predict real-world social functioning dimensionally—after accounting for diagnosis (SZ=1, HC=0), general cognition (BACS), and social cognition (MSCEIT), which are known predictors of social functioning ^84–92^. We ran Bayesian linear regressions using brms ^93^, using standardized predictors with weakly informative priors (see Supplement 20 for complete details). After controlling for diagnosis, BACS, and MSCEIT, we added DDM parameters individually and assessed whether: 1) it was a credible predictor (based on 90% HDI) and 2) it improved out-of-sample predictive accuracy (based on LOO-ELPD procedure described above). The strength of the results was interrogated using BFs. Model BFs (BF*Model*)—calculated using ‘bayes_factor’ in ^93^—indexed the strength of evidence for the full (alternative) relative to the reduced (null) model. Predictor-level BFs (BF*Predictor*)—calculated using the Savage-Dickey Density Ratio method ^94^—indexed the relative likelihood of the alternative (predictor value is not zero) relative to the null (predictor value is zero). To assess whether results were robust to antipsychotic effects, sensitivity analyses controlled for antipsychotic dosing before DDM parameters were entered. To determine whether results were specific to DDM parameters, we tested whether traditional metrics could also predict social functioning above and beyond diagnosis, BACS, and MSCEIT.

## Results

Sample characteristics are shown in Table 1.

**Table 1.**
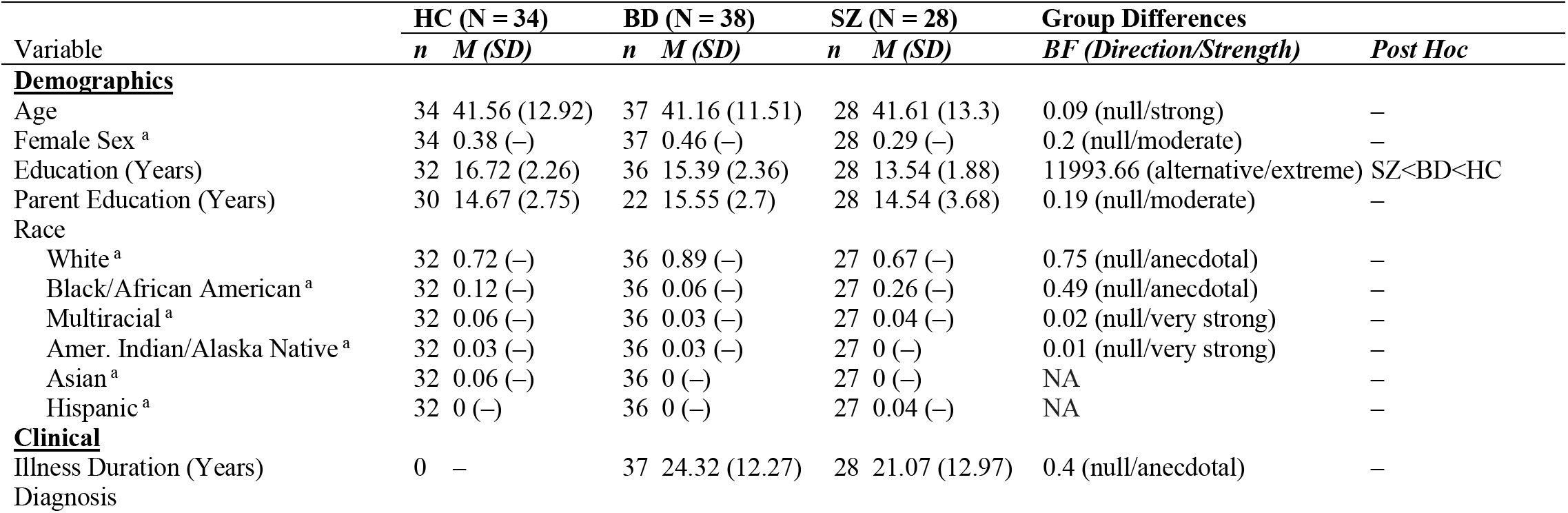
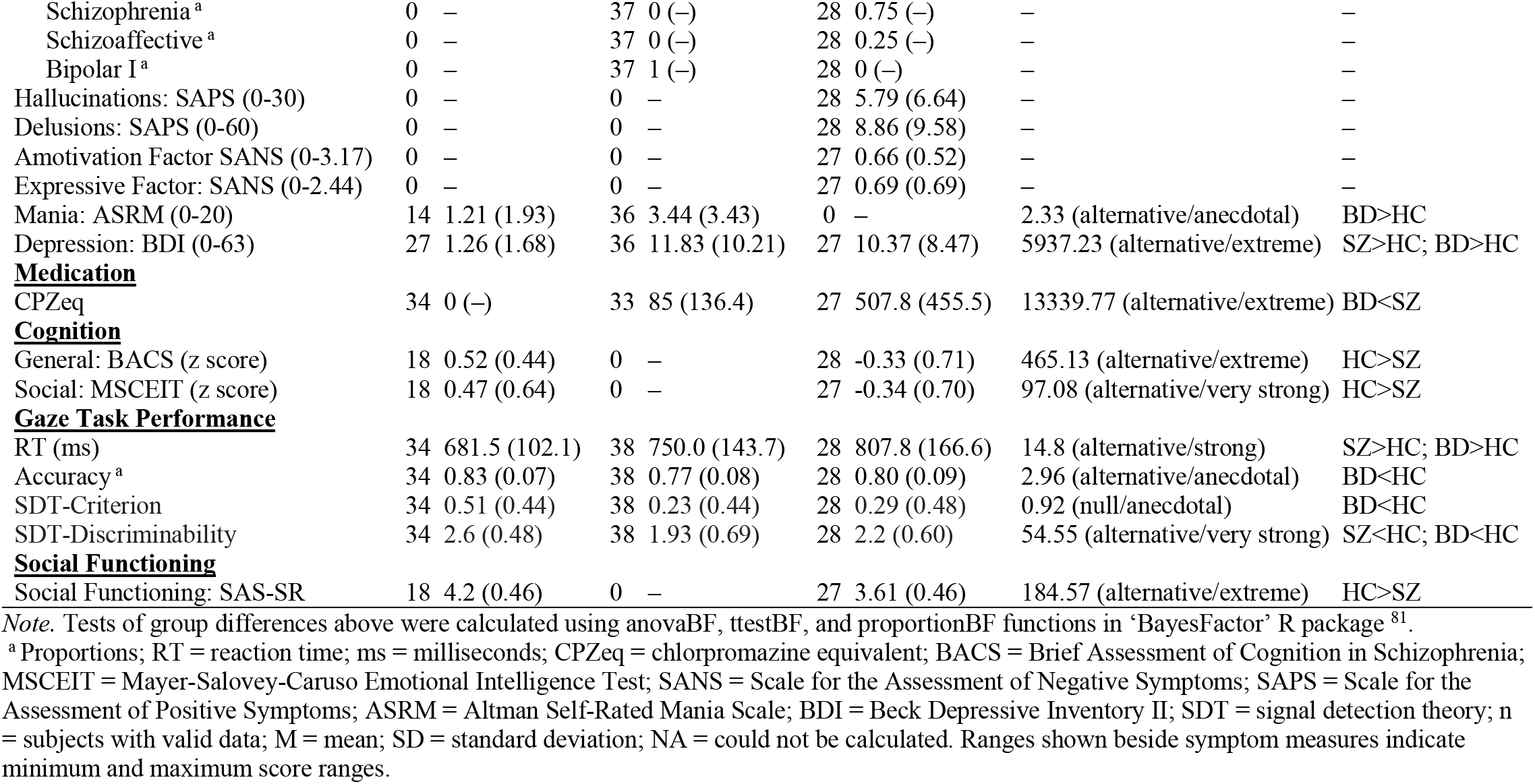
Sample Characteristics.

| Variable | HC (N = 34) |  | BD (N = 38) |  | SZ (N = 28) |  | Group Differences |  |
| --- | --- | --- | --- | --- | --- | --- | --- | --- |
|  | *n* | *M (SD)* | *n* | *M (SD)* | *n* | *M (SD)* | *BF (Direction/Strength)* | *Post Hoc* |
| **<u>Demographics</u>** |  |  |  |  |  |  |  |  |
| Age | 34 | 41.56 (12.92) | 37 | 41.16 (11.51) | 28 | 41.61 (13.3) | 0.09 (null/strong) | – |
| Female Sex<sup>a</sup> | 34 | 0.38 (–) | 37 | 0.46 (–) | 28 | 0.29 (–) | 0.2 (null/moderate) | – |
| Education (Years) | 32 | 16.72 (2.26) | 36 | 15.39 (2.36) | 28 | 13.54 (1.88) | 11993.66 (alternative/extreme) | SZ<BD<HC |
| Parent Education (Years) | 30 | 14.67 (2.75) | 22 | 15.55 (2.7) | 28 | 14.54 (3.68) | 0.19 (null/moderate) | – |
| Race |  |  |  |  |  |  |  |  |
| White<sup>a</sup> | 32 | 0.72 (–) | 36 | 0.89 (–) | 27 | 0.67 (–) | 0.75 (null/anecdotal) | – |
| Black/African American<sup>a</sup> | 32 | 0.12 (–) | 36 | 0.06 (–) | 27 | 0.26 (–) | 0.49 (null/anecdotal) | – |
| Multiracial<sup>a</sup> | 32 | 0.06 (–) | 36 | 0.03 (–) | 27 | 0.04 (–) | 0.02 (null/very strong) | – |
| Amer. Indian/Alaska Native<sup>a</sup> | 32 | 0.03 (–) | 36 | 0.03 (–) | 27 | 0 (–) | 0.01 (null/very strong) | – |
| Asian<sup>a</sup> | 32 | 0.06 (–) | 36 | 0 (–) | 27 | 0 (–) | NA | – |
| Hispanic<sup>a</sup> | 32 | 0 (–) | 36 | 0 (–) | 27 | 0.04 (–) | NA | – |
| **<u>Clinical</u>** |  |  |  |  |  |  |  |  |
| Illness Duration (Years) | 0 | – | 37 | 24.32 (12.27) | 28 | 21.07 (12.97) | 0.4 (null/anecdotal) | – |
| Diagnosis |  |  |  |  |  |  |  |  |
| Schizophrenia <sup>a</sup> | 0 | – | 37 | 0 (–) | 28 | 0.75 (–) | – | – |
| Schizoaffective <sup>a</sup> | 0 | – | 37 | 0 (–) | 28 | 0.25 (–) | – | – |
| Bipolar I <sup>a</sup> | 0 | – | 37 | 1 (–) | 28 | 0 (–) | – | – |
| Hallucinations: SAPS (0-30) | 0 | – | 0 | – | 28 | 5.79 (6.64) | – | – |
| Delusions: SAPS (0-60) | 0 | – | 0 | – | 28 | 8.86 (9.58) | – | – |
| Amotivation Factor: SANS (0-3.17) | 0 | – | 0 | – | 27 | 0.66 (0.52) | – | – |
| Expressive Factor: SANS (0-2.44) | 0 | – | 0 | – | 27 | 0.69 (0.69) | – | – |
| Mania: ASRM (0-20) | 14 | 1.21 (1.93) | 36 | 3.44 (3.43) | 0 | – | 2.33 (alternative/anecdotal) | BD>HC |
| Depression: BDI (0-63) | 27 | 1.26 (1.68) | 36 | 11.83 (10.21) | 27 | 10.37 (8.47) | 5937.23 (alternative/extreme) | SZ>HC; BD>HC |
| **<u>Medication</u>** |  |  |  |  |  |  |  |  |
| CPZeq | 34 | 0 (–) | 33 | 85 (136.4) | 27 | 507.8 (455.5) | 13339.77 (alternative/extreme) | BD<SZ |
| **<u>Cognition</u>** |  |  |  |  |  |  |  |  |
| General: BACS (z score) | 18 | 0.52 (0.44) | 0 | – | 28 | -0.33 (0.71) | 465.13 (alternative/extreme) | HC>SZ |
| Social: MSCEIT (z score) | 18 | 0.47 (0.64) | 0 | – | 27 | -0.34 (0.70) | 97.08 (alternative/very strong) | HC>SZ |
| **<u>Gaze Task Performance</u>** |  |  |  |  |  |  |  |  |
| RT (ms) | 34 | 681.5 (102.1) | 38 | 750.0 (143.7) | 28 | 807.8 (166.6) | 14.8 (alternative/strong) | SZ>HC; BD>HC |
| Accuracy <sup>a</sup> | 34 | 0.83 (0.07) | 38 | 0.77 (0.08) | 28 | 0.80 (0.09) | 2.96 (alternative/anecdotal) | BD<HC |
| SDT-Criterion | 34 | 0.51 (0.44) | 38 | 0.23 (0.44) | 28 | 0.29 (0.48) | 0.92 (null/anecdotal) | BD<HC |
| SDT-Discriminability | 34 | 2.6 (0.48) | 38 | 1.93 (0.69) | 28 | 2.2 (0.60) | 54.55 (alternative/very strong) | SZ<HC; BD<HC |
| **<u>Social Functioning</u>** |  |  |  |  |  |  |  |  |
| Social Functioning: SAS-SR | 18 | 4.2 (0.46) | 0 | – | 27 | 3.61 (0.46) | 184.57 (alternative/extreme) | HC>SZ |

*Note.* Tests of group differences above were calculated using `anovaBF`, `ttestBF`, and `proportionBF` functions in ‘BayesFactor’ R package <sup>81</sup>.
<sup>a</sup> Proportions; RT = reaction time; ms = milliseconds; CPZeq = chlorpromazine equivalent; BACS = Brief Assessment of Cognition in Schizophrenia; MSCEIT = Mayer-Salovey-Caruso Emotional Intelligence Test; SANS = Scale for the Assessment of Negative Symptoms; SAPS = Scale for the Assessment of Positive Symptoms; ASRM = Altman Self-Rated Mania Scale; BDI = Beck Depressive Inventory II; SDT = signal detection theory; n = subjects with valid data; M = mean; SD = standard deviation; NA = could not be calculated. Ranges shown beside symptom measures indicate minimum and maximum score ranges.

### Group differences in DDM parameters

Figure 3A-G plots the posterior estimates of the DDM parameters. We discuss each of these parameters in turn, starting with the parameters that most directly shape the accumulated evidence: drift rate and threshold separation.

**Figure 3.**
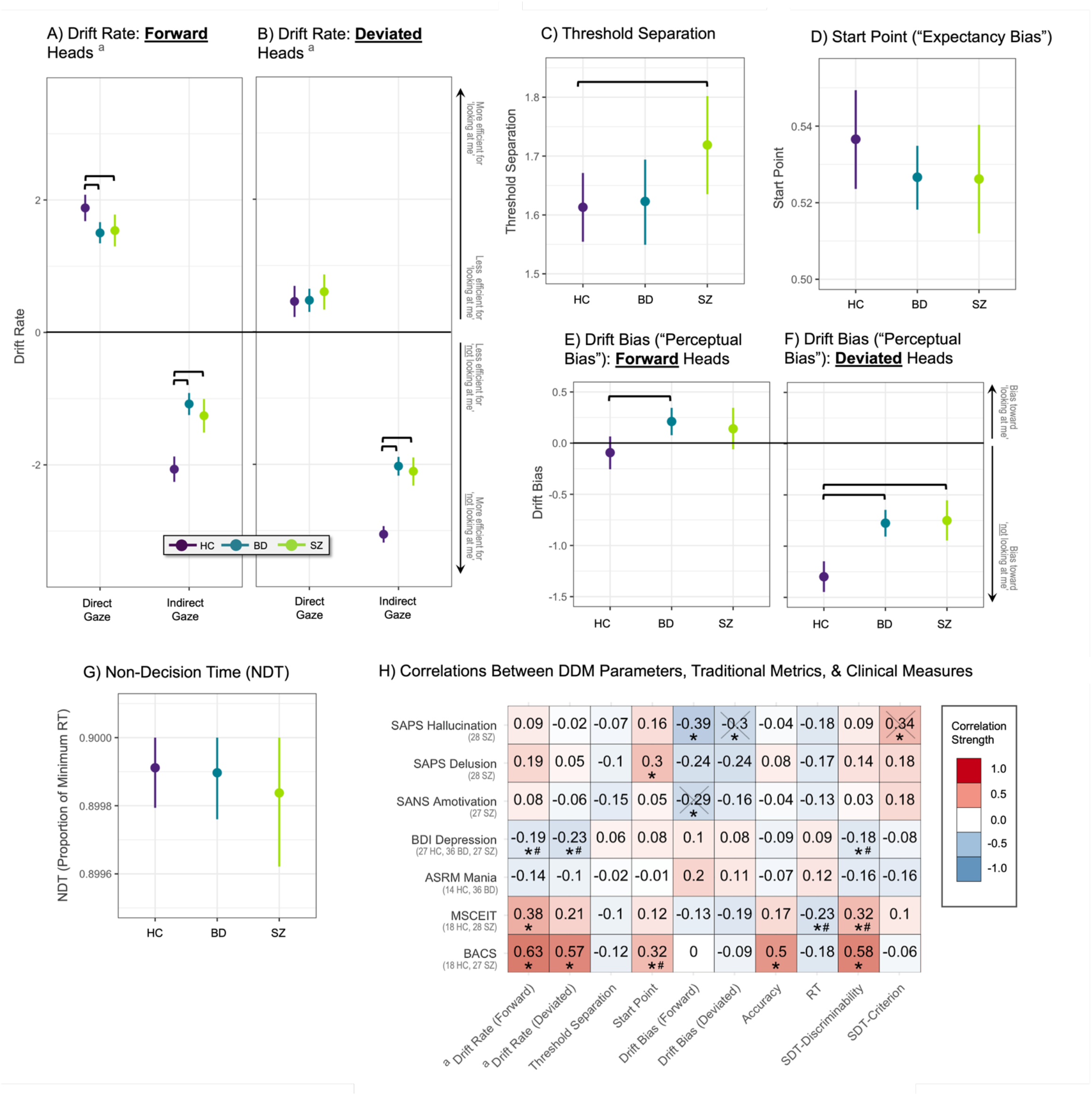
A-G) Group differences in DDM parameters. H) Zero-order correlations between DDM parameters, traditional performance metrics, general/social cognition, and SZ symptoms. *Panels A-G:* Points and error bars represent the mean and 90% highest density interval (HDI) of group posteriors, respectively. Horizontal black brackets indicate credible differences that do not contain zero. *Panel G:* NDT is a proportion of the minimum RT. *Panel H:* Values shown are mean posterior estimates of correlation coefficients (equivalent to Pearson R). The number of participants with valid data for each clinical metric is noted in the row labels. * = Credible zero-order association (i.e., 90% HDI does not contain zero); X = zero-order associations that were credible in the full sample, but were not robust to removal of influential cases; # = zero-order associations that were credible in the full sample, but were not also credible within SZ and/or BD patient groups; BACS = brief assessment of cognition in schizophrenia; DDM = drift diffusion model; MSCEIT = Mayer-Salovey-Caruso Emotional Intelligence Test; SANS = Scale for the Assessment of Negative Symptoms; SAPS = Scale for the Assessment of Positive Symptoms; BDI = Beck Depression Inventory; ASRM = Altman Self-Rated Mania Scale; RT = reaction time. ^a^ For panels A and B, drift rates were coded as follows: for direct gaze, *higher* drift rates = more efficient evidence accumulation; for indirect gaze, *lower* drift rates = more efficient evidence accumulation. For panel H, drift rates—for forward and deviated heads—were calculated such that higher values = more efficient evidence accumulation.

#### Drift rates

Recall the best-fitting model allowed the drift rate to vary by the gaze direction, head orientation, and emotional expression of stimuli. Figure 3A plots drift rates for direct and indirect gaze for forward heads, averaging across emotion conditions, while Figure 3B plots those for deviated heads (Supplement 14.2 reports group differences *before* marginalizing over emotion). As the choice option of “looking at me” was coded as the top choice threshold and “not looking at me” was coded as the bottom threshold, the drift rates reflect this coding: direct gaze faces have positive drift rates and indirect gaze faces have negative drift rates.

Overall, the drift rates indicated a general inefficiency of evidence accumulation during gaze perception in patients. This was evidenced by stronger drift rates in HC relative to SZ (M = 0.49, [0.31, 0.67]) and BD (M = 0.59, [0.45, 0.74]) when we collapsed across all task conditions. BD and SZ did not differ (M = −0.11, [−0.27, 0.06]).

Within task conditions, the pattern of results was similar. When gaze was direct and head orientation was forward, HC showed stronger drift rates than SZ (i.e., higher values in HC; M = 0.34, [0.03, 0.65]) and BD (M = 0.38, [0.12, 0.63]), while BD and SZ did not differ (M = −0.04, [−0.32, 0.26]). When gaze was indirect and head orientation was forward, HC showed stronger drift rates than SZ (i.e., lower values in HC; M = −0.81, [−1.13, −0.49]) and BD (M = −0.99, [−1.24, −0.73]), while BD and SZ did not differ (M = 0.18, [−0.13, 0.48]). When gaze was indirect and head orientation deviated, HC showed stronger drift rates than SZ (i.e., lower values in HC; M = −0.95, [−1.19, −0.70]) and BD (M = −1.03, [−1.22, −0.84]), while BD and SZ did not differ (M = 0.08, [−0.18, 0.33]). The exception was when gaze was direct and head orientation was deviated, for which all groups showed collectively poor accuracy. In this condition, there were no credible group differences for drift rates (HC-SZ: M = −0.15, [−0.51, 0.20]; HC-BD: M = - 0.02, [−0.31, 0.28]; BD-SZ: M = −0.13, [−0.45, 0.19]).

#### Threshold separation

SZ had credibly higher threshold separation than HC (Figure 3C; M = 0.11, [0.21, 0.004],) while BD did not show credible differences relative to SZ (M = −0.1, [−0.21, 0.02]) or HC (M = −0.01, [−0.10, 0.09]). This difference indicates that SZ had greater response caution than HC. This increased threshold separation helped offset the reduction in performance from decreased drift rates and may have been used in SZ as compensatory response caution, preserving accuracy at the expense of even slower RT.

#### Start point

All three groups had start points biased toward “looking at me” responses (i.e., all start points > 0.5), but there were no credible differences between groups in terms of the start points (Figure 3D; M = 0.01, HC-SZ: [−0.01, 0.03]; M = 0.01, HC-BD: [−0.01, 0.03]; BD-SZ: M = 0.00, [−0.03, 0.02]). This lack of credible differences suggests that expectancy biases are a normative aspect of gaze perception but not a driver of altered performance in SZ and BD relative to HC.

#### Drift bias

The DDM captures a second way that participants can be biased in their responses: via an asymmetry between the magnitudes of the drift rates for direct and indirect gazes. To identify this drift bias, we calculated the relative magnitudes of the drift rates for direct and indirect gaze for each condition (Figure 3E and 3F). BD showed greater drift bias towards “looking at me” responses than HC for forward (HC-BD: M = −0.30, [−0.51, −0.10]) and deviated heads (HC-BD: M = −0.523, [−0.72, −0.33]). SZ showed greater drift bias towards “looking at me” for deviated heads (HC-SZ: [−0.80, −0.31], M = −0.55), but not forward heads (HC-SZ: M = −0.23, [−0.49, 0.03]). Drift biases in SZ and BD did not differ (*Forward:* BD-SZ: M = 0.07, [−0.18, 0.31]; *Deviated:* BD-SZ: M = −0.03, [−0.26, 0.21]). These differences suggest that self-referential biases in patients are isolated to disruptions in the evidence accumulation process. This pattern of results also showed that head orientation is a predominant cue impacting perceptual biases.

Across groups, participants showed negligible-to-no drift bias toward ‘looking at me’ for forward heads (*All groups:* M = 0.09, [−0.01, 0.18]); as noted above, this bias was most evident in BD versus HC) and marked biases toward ‘not looking at me’ for deviated heads (*All groups:* M = −0.94, [-1.04, −0.85]; as noted above, this was less evident in SZ and BD versus HC).

#### Non-decision time

The groups did not show credible differences in NDT parameters, suggesting comparable time spent on sensory and motor processes (Figure 3G; HC-SZ: M = 0.0001, [−0.0002, 0.0004]; HC-BD: M = 0, [−0.0002, 0.0002]; BD-SZ: M = 0.0001, [−0.0002, 0.0004]).

### Relationships between DDM parameters, BACS, MSCEIT, and Symptoms

Figure 3H summarizes primary correlations. Supplement 19 contains complete results for all correlations, including post hoc and sensitivity analyses.

#### Relationships with BACS and MSCEIT

BACS was credibly associated with start point (r = .32 [.12, .53]), BF = 5.65), drift rates in forward (r = .63 [.50, .77], BF = 103614.43) and deviated head conditions (r = .57 [.42, .73], BF = 6314), accuracy (r = .50, [.31, .67], BF = 477.78), and SDT-Discriminability (r = .58, [.44, .74], BF = 10708.55), but not other DDM parameters or traditional metrics. Credible associations were supported by moderate to extreme BF evidence. This suggests that those with better cognitive ability tended to show more biased expectations about gaze being self-directed, more efficient evidence accumulation, and increased accuracy and perceptual sensitivity during gaze discrimination. When examined within SZ, these associations remained credible, except for the one between start point and BACS (i.e., the direction was preserved but it was not credible; Supplement 19.3.1).

MSCEIT was credibly associated with drift rate for forward heads (r = .38 [.19, .57], BF = 17.79), RT (r = -.23 [-.46, -.01], BF = 1.40), and SDT-Discriminability (r = 0.32 [.13, .55], BF = 6.19), but not other DDM parameters or traditional metrics. Evidence for these associations was anecdotal to strong. This indicates that improved emotion-based social cognition was associated with more efficient evidence accumulation (for forward heads) and faster response speed and perceptual sensitivity during gaze perception. However, when examined within SZ, only the relationship between MSCEIT and drift rate (forward) remained credible (see Supplement 19.3.1 for all associations within SZ).

Post-hoc analyses (Supplement 19.3.4) showed that performance-based metrics were not credibly associated with antipsychotic dose, suggesting that observed results were unlikely to be accounted for by antipsychotic effects.

#### Relationships with SZ symptoms

Within SZ, SAPS-Delusion scores were credibly associated with start point parameters (Figure 3H; r = .35 [.11,.59], BF = 2.26), but not other DDM parameters or traditional metrics, meaning that individuals with more delusional symptoms tended to have more biased expectations about gaze being self-directed. This association was initially supported by anecdotal evidence, but sensitivity analyses (Supplement 19.2) showed that evidence for this association strengthened (to moderate) when influential cases were removed. This also suggests it is not an artifact of extreme values. A post hoc analysis examining this relationship specifically within paranoid delusions is available in Supplement 19.3.2.

Within SZ, SAPS-Hallucination was credibly associated with drift bias for forward (r = - .39, [-.64, -.16], BF = 6.08) and deviated heads (r = -.31 [-.56, -.05], BF = 2.29), as well as SDT-Criterion (r = .35 [.11, .59], BF = 3.97). Evidence for these associations was anecdotal to moderate. This initially suggested that individuals with more severe hallucinations were *less* perceptually biased toward “looking at me” based on drift bias and SDT measures. However, sensitivity analyses (Supplement 19.2) showed that, when potentially influential cases were removed, evidence for the relationship between hallucination and drift bias for *forward* heads strengthened (from moderate to strong), but relationships with SDT-Criterion and drift bias for *deviated* heads weakened and were no longer credible. Thus, the former is more robust and the only one we will interpret.

Within SZ, SANS-Amotivation initially showed a weak credible association with drift bias for forward heads (r = -.30 [-.56, -.03], BF = 1.87), but not other DDM parameters or traditional metrics. However, it was not robust to removing potentially influential cases (Supplement 19.2).

#### Relationships with mood symptoms

Across SZ, BD, and HC, higher BDI-Depression was credibly associated with lower drift rates for forward (r = -.18 [-.35, -.03], BF = 1.29) *and* deviated heads (r = -.23 [-.39, -.07], BF = 3.21), but not other DDM parameters or traditional metrics. These were supported by anecdotal to moderate evidence. However, these associations were not evident within BD or SZ (Supplement 19.3.1-19.3.2), suggesting the relationship may have been influenced by group differences. No credible associations with ASRM-Mania emerged.

### Predicting social functioning in SZ and HC

To understand whether DDM parameters could predict real-world social functioning dimensionally (across SZ and HC), beyond well-documented predictors (BACS, MSCEIT), we performed Bayesian regression analyses (Table 2). After controlling for diagnosis, BACS, and MSCEIT, start point was a credible predictor (r = −0.3 [−0.52, −0.07], BF*Predictor* = 1.54) of real-world social functioning dimensionally across SZ and HC. Relative to the reduced model, it improved the predictive accuracy of social functioning (ΔELPD = 2.02 > ΔELPD SE = 1.99, BF*Model* = 1.55). This suggests that individuals with stronger self-referential expectation biases tended to have poorer real-world social functioning. However, BF evidence at the predictor and model level were anecdotal and should be interpreted cautiously. Although the drift rate for deviated heads was another credible predictor of social functioning (r = 0.25 [0.03, 0.46]), we do not interpret it because: it weakly improved the predictive accuracy (ΔELPD = 1.10 < ΔELPD SE = 1.42); and its inclusion was not favored by model- or predictor-level BFs (both BFs < 1).

**Table 2.** Predicting Social Functioning using Gaze DDM Parameters.

| Model/Predictor | Model Change |  |  | Predictors |  |
| --- | --- | --- | --- | --- | --- |
| | *LOO ELPD*<br>(SE) | Δ*LOO*<br>*ELPD* (SE) | *BF* (Direction/Strength) | *Mean [90% HDI]* | *BF* (Direction/Strength) |
| *Model 1* | -59.16 (3.94) | – | – |  |  |
| (Intercept) |  |  |  | 0 [-0.21, 0.23] | 0.13 (null/moderate) |
| SZ Diagnosis |  |  |  | -0.5 [-0.77, -0.22]* | 12.99 (alternative/strong) |
| BACS |  |  |  | 0.1 [-0.17, 0.37] | 0.19 (null/moderate) |
| MSCEIT |  |  |  | -0.01 [-0.27, 0.25] | 0.16 (null/moderate) |
| *Model 2<sup>a</sup>* | -59.66 (4.08) | -0.50 (0.85) | 0.27 (null/moderate) |  |  |
| (Intercept) |  |  |  | 0 [-0.21, 0.22] | 0.13 (null/moderate) |
| SZ Diagnosis |  |  |  | -0.53 [-0.81, -0.24]* | 15.06 (alternative/strong) |
| BACS |  |  |  | 0.18 [-0.12, 0.51] | 0.31 (null/moderate) |
| MSCEIT |  |  |  | 0 [-0.28, 0.25] | 0.16 (null/moderate) |
| Drift Rate (Forward) |  |  |  | -0.16 [-0.47, 0.15] | 0.27 (null/moderate) |
| *Model 3<sup>a</sup>* | -59.12 (3.73) | 0.03 (1.15) | 0.36 (null/anecdotal) |  |  |
| (Intercept) |  |  |  | 0 [-0.23, 0.21] | 0.13 (null/moderate) |
| SZ Diagnosis |  |  |  | -0.54 [-0.82, -0.27]* | 21.49 (alternative/strong) |
| BACS |  |  |  | 0.22 [-0.1, 0.55] | 0.38 (null/anecdotal) |
| MSCEIT |  |  |  | -0.05 [-0.31, 0.22] | 0.17 (null/moderate) |
| Drift Rate (Deviated) |  |  |  | -0.21 [-0.51, 0.07] | 0.36 (null/anecdotal) |
| *Model 4<sup>a</sup>* | -60.03 (4.02) | -0.88 (0.65) | 0.16 (null/moderate) |  |  |
| (Intercept) |  |  |  | 0 [-0.21, 0.22] | 0.13 (null/moderate) |
| SZ Diagnosis |  |  |  | -0.51 [-0.77, -0.22]* | 14.06 (alternative/strong) |
| BACS |  |  |  | 0.08 [-0.2, 0.36] | 0.19 (null/moderate) |
| MSCEIT |  |  |  | 0 [-0.28, 0.27] | 0.16 (null/moderate) |
| Drift Bias (Forward) |  |  |  | 0.08 [-0.14, 0.31] | 0.16 (null/moderate) |
| *Model 5<sup>a</sup>* | -58.06 (4.24) | 1.10 (1.42) | 0.75 (null/anecdotal) |  |  |
| (Intercept) |  |  |  | 0 [-0.22, 0.2] | 0.13 (null/moderate) |
| SZ Diagnosis |  |  |  | -0.54 [-0.83, -0.29]* | 22.74 (alternative/strong) |
| BACS |  |  |  | 0.08 [-0.2, 0.34] | 0.18 (null/moderate) |
| MSCEIT |  |  |  | 0.02 [-0.24, 0.28] | 0.16 (null/moderate) |
| Drift Bias (Deviated) |  |  |  | 0.25 [0.03, 0.46]* | 0.76 (null/anecdotal) |
| *Model 6<sup>a</sup>* | -57.14 (3.39) | 2.02 (1.99) | 1.55 (moderate/anecdotal) |  |  |
| (Intercept) |  |  |  | 0 [-0.2, 0.2] | 0.12 (null/moderate) |
| SZ Diagnosis |  |  |  | -0.51 [-0.78, -0.25]* | 17.98 (alternative/strong) |
| BACS |  |  |  | 0.2 [-0.07, 0.47] | 0.36 (null/anecdotal) |
| MSCEIT |  |  |  | -0.03 [-0.28, 0.22] | 0.15 (null/moderate) |
| Start Point |  |  |  | -0.3 [-0.52, -0.07]* | 1.54 (alternative/anecdotal) |
| *Model 7<sup>a</sup>* | -60.26 (3.90) | -1.10 (0.26) | 0.14 (null/moderate) |  |  |
| (Intercept) |  |  |  | 0 [-0.21, 0.22] | 0.13 (null/moderate) |
| SZ Diagnosis |  |  |  | -0.5 [-0.79, -0.23]* | 9.62 (alternative/moderate) |
| BACS |  |  |  | 0.1 [-0.19, 0.36] | 0.2 (null/moderate) |
| MSCEIT |  |  |  | -0.01 [-0.3, 0.25] | 0.16 (null/moderate) |
| Threshold Separation |  |  |  | -0.01 [-0.24, 0.22] | 0.13 (null/moderate) |

Note. \* = Credible predictor (i.e., HDI does not contain zero). BF = Bayes Factor. <sup>a</sup> Relative to Model 1. Shaded rows indicate model in which DDM parameter was a credible predictor and the change in predictive accuracy ( $\Delta$ LOO ELPD) exceeded the SE of the ELPD difference.

Post-hoc analyses (Supplement 20.2-20.3) showed that regression results were unchanged when we controlled for antipsychotic dose and were unique to DDM parameters (i.e., no traditional metrics were credible predictors or increased predictive accuracy). When regressions were performed separately within HC and SZ, the direction of relationships between start point predictors and outcomes were preserved, but they were no longer credible. This is unsurprising given the reduced sample size.

## Discussion

Self-referential gaze perception is a key social cognitive process. This paper used the DDM to delineate the precise decision-making deficits underlying altered performance in SZ and BD previously identified in the literature. Across participants, efficiency of evidence accumulation was influenced by gaze direction, head orientation, and emotion of stimuli. We uncovered process-level differences in SZ and BD, including less efficient evidence accumulation, perceptual biases predisposing self-referential responses (most prominent in BD), and greater caution (SZ only).

Patients accumulated evidence less efficiently than HC and, across SZ and HC, more efficient evidence accumulation was related to better general and emotion-based social cognition. This aligns with data ^38–41,44,95–97^ suggesting that patients’ performance alterations across tasks reflect a “general inefficiency” —potentially due to general cognitive deficits ^44^— in the ability to extract information and accumulate it as evidence to make a decision. Crucially, our results show these deficits also impact key social cognitive processes like gaze perception. This represents one avenue via which a general processing inefficiency may influence the social lives of patients with SZ and BD. Real-world social interactions require rapid processing of social cues. Inefficient evidence accumulation for such cues may disrupt the natural flow of interaction, making the individual prone to mistakes and slower processing. Although we did not find relationships between *global* social functioning and evidence accumulation, studies should explore how evidence accumulation impacts moment-to-moment interactions dynamically using performance-based assessments.

As indicated by start points, all groups showed expectancy biases toward beliefs that others were “looking at me.” This is consistent with nonclinical data showing that our default response is to endorse gaze as self-referential ^98^. Although start points did not vary between groups, they were uniquely related to functioning and delusions. *First*, across SZ and HC, more self-referential start points predicted poorer social functioning beyond diagnosis and general/social cognition. *Second*, within SZ, more self-referential start points were related to more severe delusions. Since delusions are thought to stem from maladaptive belief-level biases ^57,58^, this points to a viable mechanism that could account for links between gaze perception and delusions ^26,28^. Because our SZ sample was relatively stable, studies should examine how this association scales with increasing symptom severity. These findings echo work emphasizing the role that belief-level social cognitive biases play in general social functioning and SZ ^99–102^.

Although studies have measured such biases using self-reports ^103^ and choice-based measures ^15,24,25,29,104,105^, computational approaches offer a novel means of measuring biased social expectations.

Instead of showing biases at the belief level, patients showed self-referential *perceptual* biases toward “looking at me”, as indicated by drift bias parameters. This difference was evident in BD in all conditions, but for deviated heads only in SZ, pointing to a more widespread self-referential propensity in BD. This is consistent with evidence of excessive social approach in BD ^106–108^, which may be driven by a processing advantage for self-referential information in BD as observed here. Future studies should use self-referential versus non-self-referential tasks to ascertain whether this represents a perceptual bias in the tendency to self-reference *specifically* or to over-endorse *generally.* The latter could contribute to excessive detection of eye contact (observed here), as well as false alarms and inhibitory control failures seen in BD across tasks, which may relate to trait impulsivity. Additionally, evidence of varying degrees of perceptual bias in both patient groups raises questions about the design of neuroplasticity-based, computerized social cognitive training, which aims to broadly improve a targeted ability through repeated computerized practice exercises ^109,110^. If such interventions target processing efficiency generally but not discrepant efficiency for different choice types, biased judgments stemming from perceptual biases will remain unchanged. Instead, individualizing training to target areas of reduced processing efficiency may lead to more noticeable reductions in perception-based biases.

In SZ, greater perceptual bias toward “*not* looking at me” (forward heads only), indexed by drift bias, was associated with more severe hallucinations. Finding a relationship between perceptual biases and symptoms thought to arise from strong biases influencing perception ^56^ is promising, but the direction is unexpected. It is likely the by-product of many complicating factors that cannot be fully parsed here. Although this should be interpreted cautiously until it can be replicated, it raises a key point. Traditionally, studies have examined how aberrant biases at the belief level exert influence on belief formation and perceptions to give rise to delusions and hallucinations, respectively. However, in our case, distinguishing biases at the belief and perceptual levels via start point and drift bias revealed relationships with separate positive symptom dimensions. Ultimately, there may be value in separating biases evident at different processing levels to better understand the complex relationships between perceptual decision-making, hallucinations, and delusions.

SZ also exhibited modest increases in response caution, indexed by higher threshold separation, relative to HC during gaze perception. This, in conjunction with inefficient evidence accumulation, contributed to slower—but unbiased and accurate—responses in SZ. For SZ, heightened response caution may have functioned as a protective factor, offsetting lower drift rates to preserve response accuracy at the expense of slower RTs. Past studies modeling SZ decision processes report mixed findings, including greater caution in SZ in some studies ^39,40^ (and one interpretation of ^95^), but not others ^38,41,96,111^. This suggests that response caution is task-dependent, which is consistent with the cognitive literature that treats response caution as a means of participants’ controlling how they trade speed for accuracy. This property makes it interesting to consider from a clinical viewpoint. In many ways, this mirrors the process of cognitive restructuring in Cognitive-Behavioral Therapy, where patients slow their thinking and challenge initial perceptions to arrive at more accurate conclusions. Similar skill building may, therefore, be relevant in the augmentation of altered social perception, though it would come at the cost of slower responses. However, with practice, this may become more automatic and natural.

Alternatively, the *absence* of adjustments in response caution in BD relative to HC signals that, unlike SZ, BD did not employ additional compensation for biased and inefficient evidence accumulation. Although BD showed similar response caution to HC, controls did not need such adjustments because they did not have decrements in evidence accumulation or prominent perceptual biases to account for, as BD did. When looking at behavior indexed with traditional metrics, this contributed to self-referentially biased and less accurate responses in BD. A prevailing theory proposes that threshold separation represents a neurocomputational mechanism via which individuals exert effortful control over impulsive responses during decision-making ^112,113^. Thus, in the present study, insufficient adjustments in response caution in BD may represent one mechanism via which trait impulsivity in BD exerts influence over social cognitive abilities.

Three additional findings warrant further discussion. First, we did not find differences in NDT, which is frequently slowed in SZ ^38,39,41,96^. This may be due to our relatively simple task: prior studies used more complex/demanding ones and NDT increases with task complexity ^41^.

Additionally, slower NDT may emerge in more symptomatic samples, as motor symptoms show state- and trait- features ^114,115^. Second, we did not observe robust relationships between DDM parameters and amotivation-related negative symptoms. This may be due to the lack of consequences in our task, which may have been ill-suited to tap into amotivation. Third, although more biased start points were associated with more delusions in SZ, paradoxically, they were also associated with *improved* cognition across SZ and HC. This may be related to the fact that patients predominated by positive symptoms (“non-deficit” schizophrenia) tend to show better cognitive ability than those predominated by negative symptoms (“deficit” schizophrenia ^116^^–119^).

These findings should be interpreted considering several limitations. First, we used a task with brief stimulus durations and assumed, like prior studies, that evidence was sampled from visual short-term memory (STM) rather than the stimulus itself. Considering STM deficits in SZ, we cannot conclude whether diminished evidence accumulation efficiency is due to deficits in the ability to extract information from the stimuli or its STM representation. Future studies should disentangle these contributions. Second, we used dichotomous stimuli (self-directed or not), but there is value in including ambiguous stimuli spanning from self-directed to averted ^15,25,29,104,105^. Future studies should expand this investigation using psychophysical tasks. Third, we did not have complete data for symptoms, cognitive abilities, and functioning across the entire sample, thus limiting our ability to ascertain whether observed relationships represent transdiagnostic features.

In summary, this study used DDMs to delineate the processes driving altered self-referential gaze perception in SZ and BD. Results revealed that diminished evidence accumulation and perceptual biases may underlie altered gaze perception in SZ and BD and that SZ—but not BD—may engage in compensatory response caution to preserve the accuracy of judgments at the expense of even slower RT. DDM parameters were related to measures of general/social cognition, positive symptoms in SZ, and social functioning across SZ and HC.

Computational modeling can, therefore, provide a more nuanced understanding of the mechanisms of social cognitive difficulties in the study of psychopathology.

## Supporting information

lasagna2024_gazeDDM_supplement

## Data Availability

Participants did not provide permissions for open-access data sharing so the raw data is not publicly available. However, to promote transparency, replicability, and reproducibility, code used for modeling and analyses, is available on GitHub (github.com/CarlyLasagna/gazeddm_sz_bd) and OSF (osf.io/x5n93/?view_only=c5e6d4bf6fbe48bebe9e28d6938b9246).

https://www.github.com/CarlyLasagna/gazeddm_sz_bd

## Acknowledgements

This material is based upon work supported by the: National Science Foundation Graduate Research Fellowship under Grant No. DGE-1841052 (to CAL); National Institute of Mental Health (R01MH122491 to IFT); Prechter Bipolar Research Program. Any opinion, findings, and conclusions or recommendations expressed in this material are those of the authors(s) and do not necessarily reflect the views of the National Science Foundation. The authors report no relevant conflicts of interest. Participants did not provide permissions for open- access data sharing, so the raw data is not publicly available. However, to promote transparency, replicability, and reproducibility, code used for modeling and analyses, is available on GitHub (github.com/CarlyLasagna/gazeddm_sz_bd) and OSF (osf.io/x5n93/?view_only=c5e6d4bf6fbe48bebe9e28d6938b9246).

