## Supplementary material for "Cognitive Mechanisms of Aberrant Self-Referential Social Perception in Psychosis and Bipolar Disorder: Insights from Computational Modeling": lasagna2024_gazeDDM_supplement: supplement_gaze_ddm_revised.html

### 1 Code Availability

**Code used to perform all modeling, simulation, and analysis
is available on OSF and GitHub:**

- OSF (https://osf.io/x5n93/?view\_only=c5e6d4bf6fbe48bebe9e28d6938b9246)
- GitHub (https://github.com/CarlyLasagna/gazeddm\_sz\_bd).

### Methods Supplement

### 2 Study Sample

Participants were recruited from the University of Michigan Prechter
Bipolar Longitudinal Study, community advertisements, and local clinics.
During recruitment, distributions of HC subsamples were age- and
sex-matched to those of the SZ and BD sub-samples. Participants had no
history of medical conditions with neurological sequelae, visual acuity
of 20/30 or better on Snellen chart, and no recent substance use
disorder (patients had no substance abuse or dependence in past year, HC
in last five years). BD met criteria for bipolar I and SZ met criteria
for schizophrenia or schizoaffective disorder. HC had no history of axis
I disorders and no first-degree relatives with bipolar or psychotic
disorders.

### 3 Data Collection

Participants gave written informed consent and were compensated for
participation. The study received approval from the Institutional Review
Board at the University of Michigan Medical School. Study procedures
included: diagnostic assessments and clinical ratings by trained
assessors, neuropsychological tests of general/social cognition,
self-reports, and a behavioral gaze discrimination task. The task was
completed during the acquisition of electroencephalography (EEG) data,
but EEG analysis is outside the scope of this paper.

### 4 Assessments

Diagnoses were confirmed by trained assessors using the Structured
Clinical Interview for DSM-IV-TR or Diagnostic Interview for Genetic
Studies. In the SZ sub-study, SZ symptoms (Scale for Assessment of
Positive Symptoms [SAPS]; Scale for Assessment of Negative Symptoms
[SANS]), depressive symptoms (Beck Depression Inventory-II [BDI-II 53]),
general cognition (Brief Assessment of Cognition in Schizophrenia
[BACS]), emotion-related social cognition (Mayer-Salovey-Caruso
Emotional Intelligence Test [MSCEIT]), and social functioning (Social
Adjustment Scale Self-Report, Social/Leisure scale [SASSR-Social];
inverse-coded) were assessed. In the BD sub-study, mania (Altman
Self-Rated Mania scale [ASRM 52]) and depressive symptoms (BDI-II) were
assessed. Inter-rater reliability was > 80% for diagnoses and
clinician-assessed ratings.

For analyses in the main text, the measures of our clinical
correlates were scored as follows:

- **BACS scoring:** BACS subtests were z-scored relative
  to published age- and gender- norms of a normative HC sample and
  averaged to obtain a composite of general cognition (Keefe, 2008).
- **MSCEIT scoring:** MSCEIT subscales were converted to
  age- and gender-corrected standard scores (Mayer, 2002), z-scored
  relative to the full sample, and averaged to obtain a composite of
  emotion-based social cognition.
- **SANS-Amotivation scoring (Negative symptom-related
  amotivation factor):** The amotivation factor was calculated by
  applying published factor loadings (Sayers, 1996) to SANS
  avolition/apathy and asociality/anhedonia items and summing those to
  obtain a composite.
- **SAPS-Delusion scoring:** Delusional symptoms were
  scored at the total summed score of delusional items on the SAPS.
- **SAPS-Hallucination scoring:** Hallucination symptoms
  were scored at the total summed score of hallucination items on the
  SAPS.
- **BDI-Depression scoring:** Depressive symptoms were
  scored as the total summed score on the BDI-II.
- **ASRM-Mania scoring:** Mania symptoms were scored as
  the total summed score on the ASRM scale.

### Modeling Supplement

### 5 Defining/Refining Model Space

The steps below detail the full process of defining and refining the
full model space explored in the current study. After completing these
steps, we were left with 8 models that underwent more comprehensive
testing: 1,2,5,6,7,8,9, and 10. Full specification of these models is
provided in the next section.

Defining Model Space

| Step / Model Type | Model | Tested | Rationale | Pars Vary by GAZE | Pars Vary by HEAD | Pars Vary by EMO |
| --- | --- | --- | --- | --- | --- | --- |
| **Step 1: Define baseline model** | | | | | | |
| ***Baseline*** | | | | | | |
|  | 1 | yes | We assumed drift rate would need to vary by the condition (GAZE) that determines accuracy and this model doesn’t allow for that. So, while this is an implausible model, it does serve as our baseline | – | – | – |
| **Step 2: Define model space for models in which 1 single parameter varies by one or several task conditions** | | | | | | |
| ***Draft Rate Vary Only*** | | | | | | |
|  | 2 | yes | Plausible account of task behavior (drift rate is sensitive to stimulus properties so it is plausible that it could vary based upon any of our task conditions) | drift rate | – | – |
|  | 9 | yes | Plausible account of behavior (drift rate is sensitive to stimulus properties so it could plausibly vary based upon any task condition) | drift rate | drift rate | – |
|  | 10 | yes | Plausible account of behavior (drift rate is sensitive to stimulus properties so it could plausibly vary based upon any task condition) | drift rate | drift rate | drift rate |
|  | 5 | yes | Plausible account of behavior (drift rate is sensitive to stimulus properties so it could plausibly vary based upon any task condition) | drift rate | – | drift rate |
|  | – | no | Plausible account of behavior (drift rate is sensitive to stimulus properties so it could plausibly vary based upon any task condition) | – | drift rate | – |
|  | – | no | Exclusion Reason #1: (same as above) | – | drift rate | drift rate |
|  | – | no | Exclusion Reason #1: (same as above) | – | – | drift rate |
| ***Threshold Separation Vary Only*** | | | | | | |
|  | – | no | Exclusion Reason #1: (same as above) | threshold separation | – | – |
|  | – | no | Exclusion Reason #1: (same as above) | threshold separation | threshold separation | – |
|  | – | no | Exclusion Reason #1: (same as above) | threshold separation | threshold separation | threshold separation |
|  | – | no | Exclusion Reason #1: (same as above) | threshold separation | – | threshold separation |
|  | – | no | Exclusion Reason #1: (same as above) | – | threshold separation | – |
|  | – | no | Exclusion Reason #1: (same as above) | – | threshold separation | threshold separation |
|  | – | no | Exclusion Reason #1: (same as above) | – | – | threshold separation |
| ***Start Point Vary Only*** | | | | | | |
|  | – | no | Exclusion Reason #1: (same as above) | start point | – | – |
|  | – | no | Exclusion Reason #1: (same as above) | start point | start point | – |
|  | – | no | Exclusion Reason #1: (same as above) | start point | start point | start point |
|  | – | no | Exclusion Reason #1: (same as above) | start point | – | start point |
|  | – | no | Exclusion Reason #1: (same as above) | – | start point | – |
|  | – | no | Exclusion Reason #1: (same as above) | – | start point | start point |
|  | – | no | Exclusion Reason #1: (same as above) | – | – | start point |
| **Step 3: For plausible models defined above, build upon those by letting one additional parameter vary by task conditions. Begin with simplest model (model 2)** | | | | | | |
| ***Building Upon Model 2*** | | | | | | |
|  | 3 | partially | Tested intially, but later removed because it violated a key assumption of this version of the DDM. Exclusion Reason #2: threshold separation and start point are assumed to be set before the onset of a stimulus and, therefore, cannot vary based upon stimulus properties that aren’t predictable based upon task design. Here, the only ‘predictable’ task condition was EMO because condition were blocked by stimulus emotions. As such, we do not pursue this model further. | drift rate, threshold separation | – | – |
|  | 4 | partially | Tested intially, but later removed because it violated a key assumption of this version of the DDM. Exclusion Reason #2: (same as above) | drift rate, threshold separation, start point | – | – |
|  | – | no | Never tested because it violated a key assumption of this version of the DDM. Exclusion Reason #2: (same as above) | drift rate, start point | – | – |
|  | 6 | yes | Retained because it allowed drift rate to vary by the condition that determines accuracy (GAZE) AND the condition that threshold separation/start point parameters are allowed to vary by doesn’t violate assumptions of this version of the DDM | drift rate | – | drift rate, threshold separation |
|  | 7 | yes | Retained because it allowed drift rate to vary by the condition that determines accuracy (GAZE) AND the condition that threshold separation/start point parameters are allowed to vary by doesn’t violate assumptions of this version of the DDM | drift rate | – | drift rate, start point |
|  | 8 | yes | Retained because it allowed drift rate to vary by the condition that determines accuracy (GAZE) AND the condition that threshold separation/start point parameters are allowed to vary by doesn’t violate assumptions of this version of the DDM | drift rate | – | drift rate, threshold separation, start point |
| **Step 4: Initial model comparisons models made 2 things clear: 1) the models ran into difficulties when start point parameters varied by emotion condition (model 7 and 8); AND 2) fit was better fit when drift rate varied by gaze AND head (model 9 and 10 vs. others). Thus, the only additional models that made sense to consider building upon were model 9 and 10. Thus 2 more complex models were considered but not tested for reasons specified below** | | | | | | |
| ***Building Upon Model 9*** | | | | | | |
|  | – | no | Exclusion Reason #3: This is a plausible account of behavior that does not violate assumptions laid out thus far. Although allowing threshold separation to vary by emotion condition in simpler models (Model 6) did outperform other models where it did not vary by emotion (Model 5 and 7), the LOO difference between models was well within the SE of the LOOIC (LOO diff = 145 to 166; LOO SE ~469). So – considering that allowing threshold separation to vary by emotion offered very little to improve the model account – we decided not to pursue testing this model further and, instead, opted to continue with earlier, more parsimonious models. | drift rate | drift rate | threshold separation |
| ***Building Upon Model 10*** | | | | | | |
|  | – | no | Exclusion Reason #3: (same as above) | drift rate | drift rate | drift rate, threshold separation |

### 6 Model Specification

After refining the model space, we were left with 8 models that
underwent more comprehensive testing: 1,2,5,6,7,8,9, and 10. We provide
graphical and descriptive specification of these models is in the
sections that follow.

For all groups h (from 1 to 3, where 1=hc, 2=bd, 3=sz), subjects i
(from 1 to 100), and trials j (from 1 to max of 512), C_ij_
indicates the choice (1=yes/upper bound, 2=no/lower bound) and
RT_ij_ indicates the reaction time in seconds. RT_ij_
is Wiener distributed
W(α_i_,β_i_,δ_i_,τ_i_,), where
α_i_ is the subject-level threshold separation
(0.1<α_i_<3.9), β_i_ is the subject-level start
point (0<β_i_<1), δ_i_ is the subject-level
drift rate (-4<δ_i_<4), and τ_i_ is the
subject-level NDT (0<τ_i_<scaled minimum reaction time
[minRT_i_] for subject i). Because the Wiener distribution in
Stan allows upper-boundary responses only, lower boundary choices were
modeled using -δ_i_ for drift rate and 1-β_i_ for
start point.

To facilitate sampling, a non-centered parameterization is used. This
means that parameters are sampled in a ‘standardized’ space and then
transformed into target distributions. For a given parameter (say,
threshold separation), the diagnostic group- (α_μh_) and
subject-level means (α_i_‘) were sampled from a normal N(0,1),
while group-level variances (α_σh_) were sampled from a
positive-bound normal distribution N(0,.2)^+^. Then, the
subject’s threshold separation parameter (α_i_) was transformed
via φ(α_μh_+α_σh_ x α_i_) x 3.9+0.1, such
that the α_i_ is informed by subject- (α_i_’) and
group-level information (α_μh_ and α_σh_), transformed
to a 0<x<1 scale using the φ standard cumulative normal
distribution function, and scaled\* to the desired range.

*\*NDT is estimated as a proportion (0<x<1) and scaled by
each subject’s minRT_i_ to bring the values into units of
seconds. To help with issues sampling the NDT (i.e., the models had
difficulty sampling on the trial in which each subject’s minimum RT
occurred), we scaled the NDT by 0.9 of the subject’s actual
minRT_i_.*

For models 2, 5,6,7,8,9, and 10, gaze direction conditions, k (from 1
to 2, where 1=direct,2=indirect) were also accounted for by different
parameters as indicated below. For models 5,6,7,8,9, and 10, head
orientation conditions, m (from 1 to 2, where 1=forward,2=deviated),
and/or emotion conditions, l (from 1 to 2, where=neutral,2=fearful), are
also accounted for by different parameters as indicated below.

#### 6.1 Model 1

**Description: Group (h) and subject (i) effects for drift rate
(delta; δ), threshold separation (alpha; α), start point (beta; β), ndt
(tau; τ). (tau; τ).**

#### 6.2 Model 2

**Description: Group (h) and subject (i) effects for drift rate
(delta; δ), threshold separation (alpha; α), start point (beta; β), NDT
(tau; τ). Gaze condition effect (k) for drift rate (delta;
δ).**

#### 6.3 Model 5

**Description: Group (h) and subject (i) effects for drift rate
(delta; δ), threshold separation (alpha; α), start point (beta; β), ndt
(tau; τ). Gaze (k) and emotion (l) condition effect for drift rate
(delta; δ).**

#### 6.4 Model 6

**Description: Group (h) and subject (i) effects for drift rate
(delta; δ), threshold separation (alpha; α), start point (beta; β), ndt
(tau; τ). Gaze (k) and emotion condition (l) effect for drift rate
(delta; δ). Emotion condition (l) effect for start point (beta;
β).**

#### 6.5 Model 7

**Description: Group (h) and subject (i) effects for drift rate
(delta; δ), threshold separation (alpha; α), start point (beta; β), ndt
(tau; τ). Gaze (k) and emotion (l) condition effect for drift rate
(delta; δ). Emotion condition (l) effect for threshold separation
(alpha; α).**

#### 6.6 Model 8

**Description: Group (h) and subject (i) effects for drift rate
(delta; δ), threshold separation (alpha; α), start point (beta; β), ndt
(tau; τ). Gaze (k) and emotion (l) condition effect for drift rate
(delta; δ). Emotion condition (l) effect for start point (beta; β) and
threshold separation alpha; α).**

#### 6.7 Model 9

**Description: Group (h) and subject (i) effects for drift rate
(delta; δ), threshold separation (alpha; α), start point (beta; β), ndt
(tau; τ). Gaze (k) and head (m) condition effect for drift rate (delta;
δ).**

#### 6.8 Model 10 (Winning Model)

**Description: Group (h) and subject (i) effects for drift rate
(delta; δ), threshold separation (alpha; α), start point (beta; β), ndt
(tau; τ). Gaze (k), emotion (l), and head (m) condition effect for drift
rate (delta; δ).**

### 7 Convergence Checks

Basic convergence diagnostics were performed for all models. This
included checking for 0 divergences, ensuring that all Rhat values were
<1.1, checking that trace plots were well-mixed, verifying that
autocorrelation was low by lag of ~30, and checking that the effective
sample size (ESS) was sufficient. Models generally performed well by
these standards, suggesting no problems with convergence. Though model 7
and model 8 (which allowed start point to vary by emotion condition) has
some trouble with sampling efficiency. We do not include full
diagnostics here for brevity. Instead, we only present these complete
diagnostics below for the final winning model (Model 10).

### 8 Parameter Recovery

Parameter recovery analyses were performed at the start of model
testing (for our baseline model) and prior to analyses (for our winning
model) to ensure we could recover parameters for this task and DDM
parameterization. Details of parameter recovery from both stages of
modeling testing are detailed below.

#### 8.1 Parameter Recovery (Model 1)

We fixed NDT at 0.2 here 1) to save on storage/computing time, and 2)
because we were not interested in individual differences in NDT based on
subject-level parameter estimates.

To begin, we selected plausible values for group-level “generating
parameter values” (threshold separation=2, start point=.5, drift
rate=-0.5). Group-level posteriors for each parameter were then defined
as normal distributions with means equal to the aforementioned values
and each with a SD of 0.1. Using these group-level parameter
distributions, we randomly sampled subject-level parameter values for 25
subjects. These “subject-level generating values” were then fed into the
“rwiener” function of the RWiener package (Wabersich &
Vandekerckhove, 2014) to simulate choice and RTs for 512 trials—the
number of trials on the current task. We fit simulated data in Stan
using 1000 warmup draws and 4000 postwarmup draws sampled over multiple
chains. Convergence checks (described previously) indicated that all
models had converged. After fitting models, we calculated the means and
95% highest density intervals (HDIs) for the estimated posteriors (from
the model fits) for group- and subject-level parameters. These steps
were repeated for each of 50 separate simulations. We then evaluated
parameter recovery in 3 ways, described below in the following 3
sections.

##### 8.1.1 Group-Level Parameters (Averaged over simulations)

First, we examined how well the group-level generating values were
recovered by simulated models fits *on average*. We averaged over
the posterior means and 95% HDIs for group-level parameters for all 50
simulations; plotted generating group-level values (red X’s in plots
below) superimposed over the averaged mean and 95% HDI (points and error
bars in figure below); and assessed how well, on average, the
group-level generating values were captured by simulated model fits.

Results below suggest that group level parameters were well-recovered
on average because 1) the generating values (red X’s in plots below)
were contained within the average 95% HDI for all parameters (error bars
in plots below), and 2) the generating values are closely aligned with
the group-level posterior mean (data points in plots below) of simulated
fits on average for all parameters.

##### 8.1.2 Group-Level Parameters (Separate for each simulation)

Second, we examined group-level parameters for the individual
simulations to determine what % of the 50 simulations successfully
recovered the group-level generating parameter values (vertical black
line in plots below). Recovery was “successful” if the 95% HDI of the
fitted group-level posterior (error bars in plots below) for a given
simulation contained the original subject-level generating value. Data
in green in the plots below indicate the group-level parameters were
recovered and data in red indicates they were not.

Results below suggest that group level generating parameters were
well-recovered because the generating values (vertical black line in
plots below) were contained within the average 95% HDI (error bars in
plots below) in 94-98% of simulations for all parameters.

##### 8.1.3 Subject-Level Parameters

Third, we examined how well subject-level generating values were
captured by simulated model fits. We determined what % of subject-level
generating parameter values were successfully recovered across the 50
simulations. Recovery was “successful” if the 95% HDI of the fitted
subject-level posterior contained the original subject-level generating
value. Data in green in the plots below indicate the subject-level
generating values were recovered and data in red indicates they were
not.

Results below suggest that subject level generating parameters were
well-recovered because the generating values (x-axis) were contained
within the average 95% HDI of fitted values (error bars in plots below)
in 94-96% of simulated participants.

#### 8.2 Parameter Recovery (Model 10)

Parameter recovery was also performed for the winning model: Model
10. We used the same simulation and fitting procedure for Model 10 as we
described above for Model 1, with a few exceptions: 1) simulated data
and model fits accounted for the effects of gaze direction, head
orientation, and facial emotion on drift rates; 2) parameter recovery
was performed separately for HC, SZ, and BD; 3) group-level generating
values were defined as normal distributions with means and SDs matching
those of each diagnostic group’s group level mean posteriors from the
“final model fit” of Model 10 described in section 10 below; and 4) for
each simulation, the simulated number of subjects matched the actual
number of subjects for each group. We evaluated parameter recovery in 3
ways, as before, which is described in the following 3 sections (broken
down by by diagnostic group and task conditions).

##### 8.2.1 Group-Level Parameters (Averaged over simulations)

First, we examined how well the group-level generating values were
recovered by simulated models fits *on average*. We averaged over
the posterior means and 95% HDIs for group-level parameters for all 50
simulations; plotted generating group-level values (red X’s in plots
below) superimposed over the averaged mean and 95% HDI (points and error
bars in figure below); and assessed how well, on average, the
group-level generating values were captured by simulated model fits.
This was done separately for each diagnostic group and task
condition.

Results below suggest that group level parameters were well-recovered
on average for SZ, BD, and HC because 1) the generating values (red X’s
in plots below) were contained within the average 95% HDI for all
parameters (error bars in plots below), and 2) the generating values are
closely aligned with the group-level posterior mean (data points in
plots below) of simulated fits on average for all parameters.

##### 8.2.2 Group-Level Parameters (Separate for each simulation)

Second, we examined group-level parameters for the individual
simulations to determine what % of the 50 simulations successfully
recovered the group-level generating parameter values (vertical black
line in plots below). Recovery was “successful” if the 95% HDI of the
fitted group-level posterior (error bars in plots below) for a given
simulation contained the original subject-level generating value. Data
in green in the plots below indicate the group-level parameters were
recovered and data in red indicates they were not. This was done
separately for each diagnostic group and task condition.

Results below suggest that group level generating parameters were
well—recovered because–across all diagnostic groups and task
conditions—the generating values (vertical black line in plots below)
were contained within the average 95% HDI (error bars in plots below) in
88-100% of simulations for all parameters.

##### 8.2.3 Subject-Level Parameters

Third, we examined how well subject-level generating values were
captured by simulated model fits. We determined what % of subject-level
generating parameter values were successfully recovered across the 50
simulations. Recovery was “successful” if the 95% HDI of the fitted
subject-level posterior contained the original subject-level generating
value. Data in green in the plots below indicate the subject-level
generating values were recovered and data in red indicates they were
not. This was done separately for each diagnostic group and task
condition.

Results below suggest that subject level generating parameters were
well-recovered because the generating values (x-axis) were contained
within the average 95% HDI of fitted values (error bars in plots below)
in 92-97% of simulated participants.

### 9 Model Comparison

We performed model comparisons for models 1, 2, 5, 6, 7, 8, 9, and 10
using leave-one-out (LOO) cross-validation (Vehtari, 2017). LOO model
comparisons were performed in the full sample and within diagnostic
groups to ensure that the full sample winning model was also the winning
model within diagnostic groups.

In the tables below, models are presented in order from best- to
worst-fitting based on the expected log pointwise predictive density,
where higher ELPD values indicate better model fit. Differences in
out-of-sample predictive accuracy were assessed for all models relative
to the best fitting model based on changes in the ELPD (∆ELPD-LOO;
‘delta\_elpd’ below) relative to the best fitting model. Uncertainty
around ∆ELPD-LOO is captured through the standard error (SE;
‘delta\_elpd\_se’ below) of the estimated pointwise ELPD differences. We
consider an absolute change in ELPD < 1 SE as weak evidence for
improved predictive accuracy. Below, “pareto\_k” columns indicate the
number of observations in acceptable (“good” and “ok”) versus
problematic ranges (“bad” and “very bad”; Vehtari, 2022). Pareto K
diagnostics flag issues with the integrity of the LOO approximation and
can also signal problems with the model itself.

For all groups, the winning model (“Model 10” hereafter) was one in
which all parameters varied by diagnostic group and evidence
accumulation (drift rate) was influenced by gaze direction, head
orientation, and emotion expression of stimuli. It assumed that response
caution (threshold separation), start point (expectancy bias), and NDT
operated as trait-level processes that did not vary in response to
stimulus changes. This is a reasonable model account because the drift
rate is influenced by the physical qualities of the stimulus. Model 10
showed improvements in predictive accuracy relative to the other models
(i.e., ∆ELPD-LOO) that far exceeded the uncertainty of those estimates
(i.e., |∆ELPD-LOO| was 5 to 105 times the SE).

#### 9.1 Model Comparison (Full Sample)

First, we performed model comparisons on the full sample N=100
combined. Results showed that models perform better when the drift rate
is allowed to vary by gaze direction (e.g., Model 2 vs 1) and head
orientation (e.g., Model 9,10 vs others). The models also perform better
when drift rate varies by gaze direction, head orientation, AND emotion
(Model 10), versus when drift rate only varies by gaze direction and
head orientation (Model 9). The difference between the best fitting
model (Model 10) and the next best fitting model (Model 9) based on the
difference in ELPD LOO (“delta\_elpd” below) was over 5x greater than the
SE of the estimate (“delta\_elpd\_se” below). So, the improvement from
Model 9 to Model 10 was considerable.

Additionally, we noted some difficulties when start point is allowed
to vary by emotion (Model 7 and 8), which have some high Pareto K
values.

Model Comparison in the Full Sample

| model | looic | looic\_se | elpd\_loo | elpd\_loo\_se | delta\_elpd | delta\_elpd\_se | pareto\_k\_good | pareto\_k\_ok | pareto\_k\_bad | pareto\_k\_verybad |
| --- | --- | --- | --- | --- | --- | --- | --- | --- | --- | --- |
| m10 | 10970.44 | 507.24 | -5485.22 | 253.62 | – | – | 50058 | 4 | 0 | 0 |
| m9 | 11344.58 | 505.15 | -5672.29 | 252.57 | -187.07 | 36.01 | 50059 | 3 | 0 | 0 |
| m6 | 20253.94 | 469.87 | -10126.97 | 234.93 | -4641.75 | 102.97 | 50059 | 3 | 0 | 0 |
| m8 | 20256.08 | 469.77 | -10128.04 | 234.89 | -4642.82 | 103.74 | 50059 | 1 | 2 | 0 |
| m7 | 20398.59 | 468.85 | -10199.29 | 234.42 | -4714.07 | 102.28 | 50060 | 1 | 1 | 0 |
| m5 | 20419.81 | 469.32 | -10209.90 | 234.66 | -4724.68 | 101.49 | 50060 | 2 | 0 | 0 |
| m2 | 20697.94 | 468.45 | -10348.97 | 234.23 | -4863.75 | 104.38 | 50061 | 1 | 0 | 0 |
| m1 | 48026.44 | 339.75 | -24013.22 | 169.88 | -18528 | 176.86 | 50062 | 0 | 0 | 0 |

#### 9.2 Model Comparison (by Group)

Second, we ensure that the model comparison results from the full
sample hold within diagnostic groups. Results within groups show the
same pattern of results: Model 10 is the best fitting model within SZ,
BD, and HC. The improvement from Model 9 to Model 10 based on changes in
LOO ELPD exceeded the SE of the differences for all groups, indicating
Model 10 offered a considerably better account of the data. Based on
these results, it was clear that Model 10 offered the best fit of the
data based on LOO model comparisons.

Model Comparison by Group

| group | model | looic | looic\_se | elpd\_loo | elpd\_loo\_se | delta\_elpd | delta\_elpd\_se | pareto\_k\_good | pareto\_k\_ok | pareto\_k\_bad | pareto\_k\_verybad |
| --- | --- | --- | --- | --- | --- | --- | --- | --- | --- | --- | --- |
| **HC** | | | | | | | | | | | |
|  | m10 | -4054.07 | 305.99 | 2027.04 | 153.00 | – | – | 17162 | 2 | 0 | 0 |
|  | m9 | -3935.78 | 305.50 | 1967.89 | 152.75 | -59.15 | 20.42 | 17163 | 1 | 0 | 0 |
|  | m6 | -203.35 | 278.45 | 101.68 | 139.22 | -1925.36 | 66.61 | 17163 | 1 | 0 | 0 |
|  | m8 | -195.64 | 278.30 | 97.82 | 139.15 | -1929.22 | 67.04 | 17163 | 0 | 1 | 0 |
|  | m5 | -168.42 | 278.44 | 84.21 | 139.22 | -1942.83 | 66.13 | 17163 | 1 | 0 | 0 |
|  | m7 | -165.45 | 278.30 | 82.73 | 139.15 | -1944.31 | 66.57 | 17163 | 1 | 0 | 0 |
|  | m2 | -72.85 | 278.26 | 36.43 | 139.13 | -1990.61 | 67.42 | 17163 | 1 | 0 | 0 |
|  | m1 | 11339.41 | 197.14 | -5669.71 | 98.57 | -7696.74 | 111.63 | 17164 | 0 | 0 | 0 |
| **BD** | | | | | | | | | | | |
|  | m10 | 8999.78 | 292.73 | -4499.89 | 146.37 | – | – | 18964 | 1 | 0 | 0 |
|  | m9 | 9153.42 | 292.11 | -4576.71 | 146.06 | -76.82 | 21.92 | 18964 | 1 | 0 | 0 |
|  | m6 | 12301.55 | 271.33 | -6150.77 | 135.67 | -1650.88 | 60.23 | 18964 | 1 | 0 | 0 |
|  | m8 | 12303.47 | 271.41 | -6151.74 | 135.71 | -1651.84 | 60.55 | 18965 | 0 | 0 | 0 |
|  | m7 | 12356.91 | 270.38 | -6178.45 | 135.19 | -1678.56 | 59.36 | 18965 | 0 | 0 | 0 |
|  | m5 | 12363.14 | 270.57 | -6181.57 | 135.28 | -1681.68 | 59.04 | 18965 | 0 | 0 | 0 |
|  | m2 | 12473.79 | 270.26 | -6236.90 | 135.13 | -1737 | 60.95 | 18965 | 0 | 0 | 0 |
|  | m1 | 20918.42 | 197.45 | -10459.21 | 98.72 | -5959.32 | 101.71 | 18965 | 0 | 0 | 0 |
| **SZ** | | | | | | | | | | | |
|  | m10 | 6024.73 | 269.36 | -3012.36 | 134.68 | – | – | 13932 | 1 | 0 | 0 |
|  | m9 | 6126.95 | 266.61 | -3063.47 | 133.30 | -51.11 | 19.98 | 13932 | 1 | 0 | 0 |
|  | m8 | 8148.24 | 255.00 | -4074.12 | 127.50 | -1061.76 | 50.9 | 13931 | 1 | 1 | 0 |
|  | m6 | 8155.74 | 255.08 | -4077.87 | 127.54 | -1065.51 | 50.28 | 13932 | 1 | 0 | 0 |
|  | m7 | 8207.13 | 254.35 | -4103.57 | 127.18 | -1091.2 | 49.96 | 13932 | 0 | 1 | 0 |
|  | m5 | 8225.09 | 254.86 | -4112.54 | 127.43 | -1100.18 | 49.31 | 13932 | 1 | 0 | 0 |
|  | m2 | 8297.00 | 253.78 | -4148.50 | 126.89 | -1136.13 | 51.25 | 13933 | 0 | 0 | 0 |
|  | m1 | 15767.32 | 187.69 | -7883.66 | 93.84 | -4871.3 | 91.14 | 13933 | 0 | 0 | 0 |

### 10 Confusion Matrix

We further evaluated whether it was possible to arbitrate between
these 8 models (1, 2, 5, 6, 7, 8, 9, and 10) by generating a confusion
matrix. This allowed us to determine whether data generated by each
model could be identified as being best fit by the model that generated
it.

We simulated 50 data sets using each of these 8 models, fit all 8
models to each simulated data set, generated LOO values for each model
fit, and identified the best fitting model for each simulated data set
(i.e., model with the lowest LOO value). Simulated data was generated
using the same procedure described above for Parameter Recovery. Because
of the computational resources this process demands, we took steps to
reduce the overall run time: 1) we simulated only N=20 subjects for each
data set; 2) we reduced the warm up/post warmup samples (500 warm up,
1000 post warm up over multiple chains); 3) we fixed NDT at 0.2; and 4)
we performed this process in HC only (i.e., HC posterior values were
used to generate simulated data).

Results are summarized below. This confusion matrix indicates the
proportion of the N=50 data sets simulated using a given model (rows)
were best fit by each of the 8 models (columns), where values in rows
sum to 1. Ideally, the diagonal elements should equal 1 and off-diagonal
elements should equal 0, meaning that the model that generated the data
always fits the data best. Results show that data generated by Model
10—the best fitting model based on LOO model comparisons—was best fit by
Model 10 in 98% of simulations. The same was true of Model 9—the second
best-fitting model based on LOO model comparisons. There was more
difficulty arbitrating between simpler models, especially Models 5, 7
and 8. It is noteworthy that for Models 7 and 8 (which allowed start
point to vary based on emotion) we also noted sampling difficulties at
other model evaluation steps. This further corroborates our decision to
not explore those models further. Together, these results indicate that
it was possible to arbitrate between these two best-fitting models,
including Model 10–the winning model based on LOO model comparisons.

### 11 Posterior Predictions

After model comparisons and confusion matrices above, it Model 10 was
the best performing model. Before selecting it as our winning model to
be used for analyses, we subjected Model 10 to additional posterior
predictive checks.

As such, Models 10 was run in cmdstanr with a larger number of
samples to use to perform posterior predictive checks (i.e., 36 chains,
2500 warmup, and 2000 post warmup draws, resulting in 72,000 post warmup
samples). Using means and SD’s of posteriors for all parameters, we used
an estimation-based method to generate predicted RT quantiles and
predicted choice proportions for all task conditions, for all diagnostic
groups.

#### 11.1 Predicted RT Quantiles (Model 10)

Results show that the predicted RT quantiles (data in pink below) for
Model 10 matched the observed data well (data in teal below). This was
true for all 3 groups in all conditions (left panel below) and when we
marginalized over the emotion condition (right panel below). The
exception was cases in which there were a lower number of trials (e.g.,
indrect/forward/neutral/acc=0). This indicates the model is capable of
making accurate predictions about RT Quantiles.

Note: Posterior predicted RT quantiles were similarly accurate when
we marginalized over the emotion condition. However, we do not include
these below for brevity.

#### 11.2 Predicted Choice % (Model 10)

Results show that the predicted proportion of “yes” responses (data
in pink below) for Model 10 matched the observed data well (data in teal
below). This was true for all 3 groups in all conditions (left panel
below) and when we marginalized over the emotion condition (right panel
below). This indicates the model is capable of making accurate
predictions about choice proportions.

### 12 Winning Model Selection

To summarize the previous model evaluation steps: \* Both in the full
sample and separate diagnostic groups, Model 10 was the winning model
based on LOO model comparisons. It fit the data considerably better than
the next best-fitting model (Model 9) based on differences in ELPD. \* A
confusion matrix indicated that it was possible to arbitrate between
Model 10 and other models. \* Model 10 showed good recovery of group- and
subject-level parameters. \* Model 10 made accurate posterior predictions
of RT quantiles and choice proportions across task conditions and
diagnostic groups.

Considering all of the previous model evaluation steps, **Model
10 was selected as the winning model**. This model allowed all
parameters to vary by diagnostic group and also allowed drift rate to
vary in response to changes in gaze direction, head orientation, and
facial emotion of stimuli. This is a plausible model account of behavior
as the drift rate is known to be sensitive to the physical features of
stimuli.

### 13 Final Fit of Winning Model

After selecting Model 10 as our winning model, we ran a “final fit”
of this model using a larger number of samples. We did this to achieve
an effective sample size (ESS) of >10,000 for all parameters, to
ensure parameters we would be interpreting were sufficiently stable. The
final fit of Model 10 was run in cmdstanr with 2,500 warm up samples and
a total of 216,000 post warm up draws (obtained over 36 chains).
Convergence checks are reported below and indicate that chains converged
to target distributions.

#### 13.1 Convergence Check (Model 10)

In the final fit of the winning model (Model 10), there were 0
divergent transitions, the ESS was >10,000 for all variables, trace
plots were well-mixed, Rhat values were close to 1 (and all >1.1),
and autocorrelation was low but a lag of ~30. Together this indicated
that posteriors were stable and chains had converged to target
distributions.

##### 13.1.1 Rhat/ESS

##### 13.1.2 Trace Plots

##### 13.1.3 Autocorrelation

Note: We originally examined all 36 chains for autocorrelation. But
here we randomly select 6 and display those (for brevity)

### Analysis Supplement

### 14 Group/Condition Effects

We examined group and condition effects using group-level posteriors
for all parameters. Between group differences were tested in a pairwise
manner by subtracting the posterior for given parameter for one group
from another. If the 90% highest density interval (HDI) of the posterior
difference did not contain zero, then the difference was considered
‘credible’. The same process was followed to evaluate the credibility of
task condition effects within groups. This approach was appropriate
because the parameterization of the models inherently accounted for the
shared group-level variability of within-group effects.

Results presented below and detail group differences in threshold
separation, start point, NDT, and drift rate parameters. Because the
winning model (Model 10) accounted for within-group condition effects on
drift rates–for different gaze, head, and emotion conditions–we also
present task condition effects on drift rate below (both in isolation
[marginalized across group] and in combination with group effects).

#### 14.1 Threshold Separation, Start Point, NDT (Group Effects)

Results show that threshold separation is credibly higher in SZ than
HC, but no other group differences are credible for threshold
separation. The three groups exhibited comparable values for start point
and NDT parameters.

#### 14.2 Drift Rate (Group/Gaze/Head/Emo Effects)

We looked at group differences in drift rates broken down by gaze and
head conditions. This was done separately by neutral and fearful
emotions. Results showed credible group differences in drift rates
within gaze and head conditions. None of the groups exhibited group
differences when gaze was direct and heads were deviated (this was true
for both fearful and neutral emotions). When heads were forward and gaze
was direct, HC had credibly higher drift rates than BD in both emotion
conditions. When heads were forward and gaze was direct, HC had credibly
higher drift rates than SZ only in the fearful emotion condition.

#### 14.3 Drift Rate (Group/Gaze/Head Effects)

These results are reported in the main text. Here, we look at group
differences in group-level drift rate parameters in a pairwise manner.
This is done separately for drift rates in gaze and head conditions,
after marginalizing over the emotion conditions.

#### 14.4 Drift Rate (Group Effects)

We marginalized over all task conditions (emo, then head, then gaze)
for drift rates to obtain a measure of overall evidence accumulation
efficiency for all three groups. Results showed that HC had credibly
higher drift rates than SZ and BD when samples were marginalized over
all task conditions.

#### 14.5 Drift Rate (Emo Effects)

We examined the influence of emotion on drift rates within gaze/head
conditions and diagnostic groups to get an overall indicator of the
effect of emotion on drift rates. Results show that there are not
credible differences in drift rates between emotion conditions (i.e.,
all of the 90% HDI’s below contain zero) in any of the groups or
head/gaze conditions. Because of this, we chose to marginalize over
emotions conditions for drift rates and drift bias to simplify the
analyses in the main text. We chose not to marginalize over gaze and
head orientation because we did find at least some credible influence of
gaze/head cues (reported below) on drift rates within groups.
Additionally, for correlation analyses, we chose to further marginalize
over gaze direction for drift rates to reduce the number of
comparisons.

**Marginalizing over emotion conditions:** Marginalizing
over emotion conditions may appear counterintuitive when the winning
model—accounting for the influences of gaze, head, and emotion on drift
rates—outperformed other models that did not also account for the
influence of emotion on drift rates. This is an open question, but it is
likely that Model 10 captured subtle nuances in how emotion influenced
evidence accumulation that helped improve out-of-sample predictions but
were not sufficiently large to yield credible condition-level effects,
when tested at the group-level as we did.

One explanation is that Model 10 better captured variability at the
subject-level and not the group-level as our tests of condition-level
effects examined. In other words, the lack of within-group differences
in drift rates based on emotion may not have occurred universally within
the subjects of each group. Rather, condition-level emotions effects may
have occurred for a subset of individuals within each group. In this
case, the added variability introduced by emotion effects for some—but
not all—subjects would have been best captured by Model 10. However, if
this occurred in just a subset of subjects, these effects would not be
sufficiently influential to produce group-level emotion effects in the
group-level hyperparameters for the drift rates.

Although we chose to marginalize over samples in the analyses in the
main text, for completeness and transparency we also report the tests of
group differences in drift rates before marginalizing over the emotion
condition in the supplement section named “Drift Rate
(Group/Gaze/Head/Emo Effects)”.

Posterior Differences in Drift Rates Between Emotion Conditions (by Dx
Group, Gaze Direction, Head Orientation)

| group | cond\_gaze | cond\_head | emo\_contrast | HDI\_lo | HDI\_hi | mean |
| --- | --- | --- | --- | --- | --- | --- |
| hc | direct | forward | neutral-fearful | -0.379 | 0.271 | -0.057 |
| bd | direct | forward | neutral-fearful | -0.266 | 0.246 | -0.014 |
| sz | direct | forward | neutral-fearful | -0.327 | 0.463 | 0.076 |
| hc | indirect | forward | neutral-fearful | -0.251 | 0.368 | 0.054 |
| bd | indirect | forward | neutral-fearful | -0.264 | 0.280 | 0.015 |
| sz | indirect | forward | neutral-fearful | -0.492 | 0.334 | -0.079 |
| hc | direct | deviated | neutral-fearful | -0.333 | 0.449 | -0.333 |
| bd | direct | deviated | neutral-fearful | -0.108 | 0.455 | 0.178 |
| sz | direct | deviated | neutral-fearful | -0.168 | 0.695 | 0.264 |
| hc | indirect | deviated | neutral-fearful | -0.229 | 0.163 | -0.031 |
| bd | indirect | deviated | neutral-fearful | -0.367 | 0.084 | -0.142 |
| sz | indirect | deviated | neutral-fearful | -0.549 | 0.131 | -0.208 |

#### 14.6 Drift Rate (Gaze Effects)

After marginalizing over emotion conditions, we examined the
influence of gaze direction on drift rates in all three groups. Results
show facilitation effects of gaze direction in all three groups for
deviated heads (higher drift rates for indirect gaze when heads are
deviated). However, we don’t see this as consistently for forwards
heads.

Posterior Differences in Drift Rates Between Gaze Direction Conditions
(by Dx Group, Head Orientation)

| group | cond\_head | gaze\_contrast | HDI\_lo | HDI\_hi | mean |
| --- | --- | --- | --- | --- | --- |
| hc | forward | direct-indirect | -0.511 | 0.130 | -0.186 |
| bd | forward | direct-indirect | 0.154 | 0.689 | 0.422 |
| sz | forward | direct-indirect | -0.120 | 0.690 | 0.281 |
| hc | deviated | direct-indirect | -2.895 | -2.295 | -2.597 |
| bd | deviated | direct-indirect | -1.814 | -1.291 | -1.551 |
| sz | deviated | direct-indirect | -1.890 | -1.105 | -1.500 |

#### 14.7 Drift Rate (Head Effects)

After marginalizing over emotion conditions, we examined the
influence of head orientation on drift rates in all three groups.
Results show credible differences in drift rate based on the head
orientation direction of stimuli, such that individuals show more
efficient evidence accumulation for forward vs deviated heads. This is
found in all three groups.

Posterior Differences in Drift Rates Between Head Orientation Conditions
(by Dx Group, Gaze Direction)

| group | cond\_gaze | head\_contrast | HDI\_lo | HDI\_hi | mean |
| --- | --- | --- | --- | --- | --- |
| hc | direct | forward-deviated | 1.163 | 1.669 | 1.422 |
| bd | direct | forward-deviated | 0.836 | 1.222 | 1.027 |
| sz | direct | forward-deviated | 0.639 | 1.224 | 0.934 |
| hc | indirect | forward-deviated | -1.177 | -0.806 | -0.989 |
| bd | indirect | forward-deviated | -1.126 | -0.768 | -0.947 |
| sz | indirect | forward-deviated | -1.115 | -0.581 | -0.847 |

#### 14.8 Drift Bias (Group/Head Effects)

These results are presented in the main text. After marginalizing
over emotion conditions, we examined the influence of diagnostic group
on drift bias within forward and deviated head orientations. Results
showed that HC had credibly lower drift bias than BD in forward and
deviated head conditions. HC only showed credibly lower drift bias than
SZ in deviated head conditions.

#### 14.9 Drift Bias (Group Effects)

We examined the influence of diagnostic group on drift bias after
marginalizing over emotion and head conditions to get an an overall
indicator of the effect of group on drift bias. Results showed that, in
general, HC had credibly lower drift bias than BD and SZ.

#### 14.10 Drift Bias (Head Effects)

We calculated 90% HDIs for drift biases within both head orientation
conditions after marginalizing over emotion conditions and groups.
Results showed that biases toward self referential choices tended to be
higher when heads were forward and lower when heads were deviated.

Drift Bias by Head Orientation Marginalized Over Group

| head | hdi\_lo | hdi\_hi | mean |
| --- | --- | --- | --- |
| forward | -0.011 | 0.183 | 0.086 |
| deviated | -1.035 | -0.849 | -0.941 |

### 15 Preprocessing Correlates

Before conducting analyses on individual differences (correlations,
regressions), we performed necessary preprocessing steps on our
variables of interest.

1. For subject-level drift rates and drift biases, we marginalized
   over emotion conditions (as was done for group-level parameters). To
   reduce the number of tests/comparisons correlations and regressions, we
   also marginalized over gaze conditions for drift rates. This was done by
   flipping the sign of drift rates for indirect gaze (which were
   originally negative-going) and then averaging over subject-level
   posteriors for direct and indirect gaze. This was done separately for
   both forward and deviated head conditions. As a result, for all
   subejcts, we had measures of drift rates and drift biases in both
   forward and deviated head conditions.
2. To retain as much data as possible, we also winsorized the
   outermost .01 of data for variables that contained outliers.

### 16 SDT Analysis

We performed additional signal detection analyses to extract measures
of sensitivity (discriminability parameter) and bias (criterion
parameter) from participants’ choices on the gaze task. To do so, we
programmed an equal variance gaussian SDT model in Stan that accounted
for effects of head orientation on participant’s discriminability and
criterion values. The SDT model was a hierarchical Bayesian model with
weakly informative priors. These were fit separately to participants in
SZ, BD, and HC groups (equivalent to how between-group effects were
programmed in DDM’s). Models were run with 1,000 warmup samples and a
total of 40,000 postwarmup samples obtained over several chains. As with
DDM’s convergence checks were performed to ensure models had converged.
There were no divergences for any model fits. Moreover, for all
parameters of all SDT models fits, Rhat values were close to 1 (all were
<1.1), trace plots were well-mixed, autocorrelation was low by a lag
of ~30, and bulk and tail ESS were >10,000. Together, this suggested
that chains had converged to their target distributions.

*Note: For brevity we do not include outputs of convergence checks
for SDT models here.*

### 17 Interpreting Bayes Factors

For all statistical tests reported below, we calculate Bayes Factors
(BF) and use those as an additional piece of information indicating the
strength of an observed result. BF values index the evidence for the
alternative hypothesis compared against that of the null hypothesis. BF
> 1 favors the alternative hypothesis and BF < 1 favors the null
hypothesis.

- For demographic group differences: alternative hypothesis =
  difference between groups is not zero; null hypothesis = difference
  between groups is zero.
- For correlations: alternative hypothesis = association is not zero;
  null hypothesis = association is zero.
- For regression model comparisons: alternative hypothesis = favors
  the full model; null hypothsis = favors the reduced model.
- For regression predictors: alternative hypothesis = value of
  predictor is not zero; null hypothesis = value of predictor is
  zero.

We then interpret each BF using the ranges below, from Lee and
Wagenmakers (2014):

Bayes Factor Interpretation Scheme from Lee and Wagenmakers (2014)

| BF Value | Evidence Favors | Strength of Evidence |
| --- | --- | --- |
| >100 | Alternative | Extreme |
| 30-100 | Alternative | Very Strong |
| 10-30 | Alternative | Strong |
| 3-10 | Alternative | Moderate |
| 1-3 | Alternative | Anecdotal |
| 1 |  | No Evidence |
| 0.33-1 | Null | Anecdotal |
| 0.1-0.33 | Null | Moderate |
| 0.03-0.1 | Null | Strong |
| 0.01-0.03 | Null | Very Strong |
| <0.01 | Null | Extreme |

### 18 Sample Demographics

Tests below are Bayesian t-tests, ANOVA, and proportion analyses run
using ttestBF, anovaBF, and proportion BF in the ‘BayesFactor’ R package
(Morey, 2022).

Sample Characteristics

|  | HC (M) | HC (SD) | HC (N) | BD (M) | BD (SD) | BD (N) | SZ (M) | SZ (SD) | HC (N) | GroupDiff | PostHoc |
| --- | --- | --- | --- | --- | --- | --- | --- | --- | --- | --- | --- |
| **Demographic** | | | | | | | | | | | |
| Age | 41.56 | 12.92 | 34 | 41.16 | 11.51 | 37 | 41.61 | 13.3 | 28 | BF=0.09 (Null/Strong) | BD≈HC; BD≈SZ; HC≈SZ |
| Sex (% Female) | 0.38 | – | 34 | 0.46 | – | 37 | 0.29 | – | 28 | BF=0.2 (Null/Moderate) | BD≈HC; BD≈SZ; HC≈SZ |
| Education (Years) | 16.72 | 2.26 | 32 | 15.39 | 2.36 | 36 | 13.54 | 1.88 | 28 | BF=11993.66 (Alternative/Extreme) | BD<HC; BD>SZ; HC>SZ |
| Parental Education (Years) | 14.67 | 2.75 | 30 | 15.55 | 2.7 | 22 | 14.54 | 3.68 | 28 | BF=0.19 (Null/Moderate) | BD≈HC; BD≈SZ; HC≈SZ |
| *Race* | | | | | | | | | | | |
| White (%) | 0.72 | – | 32 | 0.89 | – | 36 | 0.67 | – | 27 | BF=0.75 (Null/Anecdotal) | BD≈HC; BD>SZ; HC≈SZ |
| Black or African American (%) | 0.12 | – | 32 | 0.06 | – | 36 | 0.26 | – | 27 | BF=0.49 (Null/Anecdotal) | BD≈HC; BD<SZ; HC≈SZ |
| Multiracial (%) | 0.06 | – | 32 | 0.03 | – | 36 | 0.04 | – | 27 | BF=0.02 (Null/Very Strong) | BD≈HC; BD≈SZ; HC≈SZ |
| American Indian or Alaska Native (%) | 0.03 | – | 32 | 0.03 | – | 36 | 0 | – | 27 | BF=0.01 (Null/Very Strong) | BD≈HC; BD≈SZ; HC≈SZ |
| Asian (%) | 0.06 | – | 32 | 0 | – | 36 | 0 | – | 27 | – | – |
| Hispanic (%) | 0 | – | 32 | 0 | – | 36 | 0.04 | – | 27 | – | – |
| **Clinical** | | | | | | | | | | | |
| Illness Duration | – | – | 0 | 24.32 | 12.27 | 37 | 21.07 | 12.97 | 28 | BF=0.4 (Null/Anecdotal) | BD≈SZ |
| *Diagnosis* | | | | | | | | | | | |
| Schizophrenia (%) | – | – | 0 | 0 | – | 37 | 0.75 | – | 28 | – | – |
| Schizoaffective (%) | – | – | 0 | 0 | – | 37 | 0.25 | – | 28 | – | – |
| Bipolar I (%) | – | – | 0 | 1 | – | 37 | 0 | – | 28 | – | – |
| *Symptoms* | | | | | | | | | | | |
| ASRM | 1.21 | 1.93 | 14 | 3.44 | 3.43 | 36 | – | – | 0 | BF=2.33 (Alternative/Anecdotal) | BD>HC |
| BDI | 1.26 | 1.68 | 27 | 11.83 | 10.21 | 36 | 10.37 | 8.47 | 27 | BF=5937.23 (Alternative/Extreme) | BD>HC; BD≈SZ; HC<SZ |
| SAPS-Hallucination | – | – | 0 | – | – | 0 | 5.79 | 6.64 | 28 | – | – |
| SAPS-Delusion | – | – | 0 | – | – | 0 | 8.86 | 9.58 | 28 | – | – |
| SANS-Motivation | – | – | 0 | – | – | 0 | 0.66 | 0.52 | 27 | – | – |
| SANS-Expressive | – | – | 0 | – | – | 0 | 0.69 | 0.69 | 27 | – | – |
| *Medications* | | | | | | | | | | | |
| Antipsychotic User (%) | – | – | 0 | 0.44 | – | 36 | 0.93 | – | 28 | – | – |
| Antidepressant User (%) | – | – | 0 | 0.56 | – | 36 | 0.32 | – | 28 | – | – |
| Mood Stabilizer User (%) | – | – | 0 | 0.72 | – | 36 | 0.18 | – | 28 | – | – |
| Stimulant User (%) | – | – | 0 | 0.18 | – | 33 | 0.04 | – | 28 | – | – |
| Hypnotic User (%) | – | – | 0 | 0.09 | – | 33 | 0.04 | – | 28 | – | – |
| Anxiolytic User (%) | – | – | 0 | 0.24 | – | 33 | 0.21 | – | 28 | – | – |
| Anticholinergic User (%) | – | – | 0 | 0 | – | 33 | 0.18 | – | 28 | – | – |
| CPZeq | – | – | 0 | 85 | 136.44 | 33 | 489.42 | 397.68 | 27 | BF=13339.77 (Alternative/Extreme) | BD<SZ |
| **General/Social Cognition** | | | | | | | | | | | |
| Cognition-General: BACS | 0.52 | 0.44 | 18 | – | – | 0 | -0.33 | 0.71 | 28 | BF=465.13 (Alternative/Extreme) | HC>SZ |
| Cognition-Social: MSCEIT | 0.47 | 0.64 | 18 | – | – | 0 | -0.34 | 0.7 | 27 | BF=97.08 (Alternative/Very Strong) | HC>SZ |
| **Gaze Task Performance** | | | | | | | | | | | |
| Gaze Task: RT (ms) | 681.46 | 102.05 | 34 | 750.01 | 143.67 | 38 | 807.82 | 166.63 | 28 | BF=14.8 (Alternative/Strong) | BD>HC; BD≈SZ; HC<SZ |
| Gaze Task: Accuracy | 0.83 | 0.07 | 34 | 0.77 | 0.08 | 38 | 0.8 | 0.09 | 28 | BF=2.96 (Alternative/Anecdotal) | BD<HC; BD≈SZ; HC≈SZ |
| Gaze Task: Criterion | 0.49 | 0.48 | 34 | 0.23 | 0.46 | 38 | 0.29 | 0.49 | 28 | BF=0.92 (Null/Anecdotal) | BD<HC; BD≈SZ; HC≈SZ |
| Gaze Task: Discriminability | 2.63 | 0.6 | 34 | 1.95 | 0.76 | 38 | 2.17 | 0.82 | 28 | BF=54.55 (Alternative/Very Strong) | BD<HC; BD≈SZ; HC>SZ |
| **Social Functioning** | | | | | | | | | | | |
| Social Functioning: SAS-SR | 4.2 | 0.46 | 18 | – | – | 0 | 3.61 | 0.46 | 27 | BF=184.57 (Alternative/Extreme) | HC>SZ |

### 19 Correlations

We ran exploratory Correlations Between DDM Parameters, traditional
performance metrics (Gaze accuracy, RT, signal detection
discriminability and criterion), general cognition measures (BACS),
emotion-based social cognition measures (MSCEIT), SZ symptoms
(SANS/SAPS), and mood symptoms (BDI/ASRM). This was done using functions
from the BayesFactor R package (Morey, 2022). For each correlation, we
calculate the mean and 90% HDI of the correlation coefficient, and the
Bayes factor (BF). We interpret associations using the 90% HDI:
intervals that do not contain zero are considered credible. For each
credible association, we also consider – as an added source of
information – the strength of the evidence using its BF.

The sections that follow present a correlation matrix plot (to
illustrate the nature of relationships between all measures);
sensitivity analyses to test the robustness of observed relationships;
and a series of post hoc follow-up tests, including correlations with SZ
and BD, as well as associations looking at paranoia in SZ
specifically.

#### 19.1 Correlations

Below is a full correlation matrix plot with correlations to
illustrate the nature of relationships between DDM parameters (in SZ,
BD, HC), traditional metrics (in SZ, BD, HC), BACS/MSCEIT (in SZ, HC),
SZ symptoms (SAPS/SANS; in SZ), depressive symptoms (BDI; in SZ, BD,
HC), and mania symptoms (ASRM; in BD, HC).

In the plot below, large text displayed in cells below represent the
posterior means of the correlation coefficient. This value is equivalent
to the Pearson R value. The magnitude and direction of the posterior
mean estimates control the color scheme of the correlation matrix
(positive-going relationships are in red and negative-going are in
blue). 90% HDI’s that do not contain zero are considered credible
(marked with a ’\*’ below).

Correlations Between DDM Parameters, Traditional Metrics, and Clinical
Metrics (Full Sample)

| Variable1 | Variable2 | Mean | HDI | BF | BF\_Evidence\_Strength | BF\_Evidence\_Favors | N |
| --- | --- | --- | --- | --- | --- | --- | --- |
| Drift Rate (Forward) | BACS | 0.63 | [0.5,0.77]\* | 103614.43 | Extreme | Alternative | 46 |
| Drift Rate (Forward) | MSCEIT | 0.38 | [0.19,0.57]\* | 17.79 | Strong | Alternative | 45 |
| Drift Rate (Forward) | ASRM Mania | -0.14 | [-0.36,0.08] | 0.56 | Anecdotal | Null | 50 |
| Drift Rate (Forward) | BDI Depression | -0.18 | [-0.35,-0.03]\* | 1.29 | Anecdotal | Alternative | 90 |
| Drift Rate (Forward) | SANS Amotivation | 0.08 | [-0.2,0.37] | 0.46 | Anecdotal | Null | 27 |
| Drift Rate (Forward) | SAPS Delusion | 0.19 | [-0.05,0.48] | 0.79 | Anecdotal | Null | 28 |
| Drift Rate (Forward) | SAPS Hallucination | 0.08 | [-0.21,0.35] | 0.46 | Anecdotal | Null | 28 |
| Drift Rate (Deviated) | BACS | 0.57 | [0.42,0.72]\* | 6314.00 | Extreme | Alternative | 46 |
| Drift Rate (Deviated) | MSCEIT | 0.21 | [-0.01,0.42] | 1.02 | Anecdotal | Alternative | 45 |
| Drift Rate (Deviated) | ASRM Mania | -0.10 | [-0.3,0.13] | 0.42 | Anecdotal | Null | 50 |
| Drift Rate (Deviated) | BDI Depression | -0.23 | [-0.39,-0.07]\* | 3.21 | Moderate | Alternative | 90 |
| Drift Rate (Deviated) | SANS Amotivation | -0.06 | [-0.36,0.22] | 0.45 | Anecdotal | Null | 27 |
| Drift Rate (Deviated) | SAPS Delusion | 0.05 | [-0.22,0.34] | 0.43 | Anecdotal | Null | 28 |
| Drift Rate (Deviated) | SAPS Hallucination | -0.03 | [-0.31,0.26] | 0.42 | Anecdotal | Null | 28 |
| Threshold Separation | BACS | -0.12 | [-0.35,0.1] | 0.47 | Anecdotal | Null | 46 |
| Threshold Separation | MSCEIT | -0.10 | [-0.33,0.13] | 0.44 | Anecdotal | Null | 45 |
| Threshold Separation | ASRM Mania | -0.02 | [-0.24,0.2] | 0.32 | Moderate | Null | 50 |
| Threshold Separation | BDI Depression | 0.06 | [-0.1,0.23] | 0.29 | Moderate | Null | 90 |
| Threshold Separation | SANS Amotivation | -0.14 | [-0.44,0.12] | 0.59 | Anecdotal | Null | 27 |
| Threshold Separation | SAPS Delusion | -0.10 | [-0.37,0.2] | 0.49 | Anecdotal | Null | 28 |
| Threshold Separation | SAPS Hallucination | -0.07 | [-0.37,0.2] | 0.45 | Anecdotal | Null | 28 |
| Start Point | BACS | 0.32 | [0.11,0.51]\* | 5.65 | Moderate | Alternative | 46 |
| Start Point | MSCEIT | 0.12 | [-0.12,0.33] | 0.48 | Anecdotal | Null | 45 |
| Start Point | ASRM Mania | -0.01 | [-0.23,0.21] | 0.32 | Moderate | Null | 50 |
| Start Point | BDI Depression | 0.08 | [-0.08,0.25] | 0.34 | Anecdotal | Null | 90 |
| Start Point | SANS Amotivation | 0.05 | [-0.24,0.33] | 0.44 | Anecdotal | Null | 27 |
| Start Point | SAPS Delusion | 0.31 | [0.05,0.56]\* | 2.26 | Anecdotal | Alternative | 28 |
| Start Point | SAPS Hallucination | 0.17 | [-0.1,0.44] | 0.67 | Anecdotal | Null | 28 |
| Drift Bias (Forward) | BACS | -0.01 | [-0.25,0.22] | 0.33 | Anecdotal | Null | 46 |
| Drift Bias (Forward) | MSCEIT | -0.13 | [-0.37,0.08] | 0.51 | Anecdotal | Null | 45 |
| Drift Bias (Forward) | ASRM Mania | 0.20 | [-0.01,0.4] | 1.00 | Anecdotal | Alternative | 50 |
| Drift Bias (Forward) | BDI Depression | 0.10 | [-0.07,0.27] | 0.39 | Anecdotal | Null | 90 |
| Drift Bias (Forward) | SANS Amotivation | -0.29 | [-0.54,-0.01]\* | 1.87 | Anecdotal | Alternative | 27 |
| Drift Bias (Forward) | SAPS Delusion | -0.25 | [-0.49,0.03] | 1.22 | Anecdotal | Alternative | 28 |
| Drift Bias (Forward) | SAPS Hallucination | -0.38 | [-0.65,-0.14]\* | 6.08 | Moderate | Alternative | 28 |
| Drift Bias (Deviated) | BACS | -0.09 | [-0.3,0.15] | 0.41 | Anecdotal | Null | 46 |
| Drift Bias (Deviated) | MSCEIT | -0.19 | [-0.41,0.03] | 0.84 | Anecdotal | Null | 45 |
| Drift Bias (Deviated) | ASRM Mania | 0.11 | [-0.12,0.31] | 0.44 | Anecdotal | Null | 50 |
| Drift Bias (Deviated) | BDI Depression | 0.08 | [-0.09,0.24] | 0.34 | Anecdotal | Null | 90 |
| Drift Bias (Deviated) | SANS Amotivation | -0.16 | [-0.43,0.12] | 0.64 | Anecdotal | Null | 27 |
| Drift Bias (Deviated) | SAPS Delusion | -0.24 | [-0.51,0.03] | 1.16 | Anecdotal | Alternative | 28 |
| Drift Bias (Deviated) | SAPS Hallucination | -0.30 | [-0.57,-0.03]\* | 2.29 | Anecdotal | Alternative | 28 |
| Accuracy | BACS | 0.50 | [0.32,0.67]\* | 477.78 | Extreme | Alternative | 46 |
| Accuracy | MSCEIT | 0.16 | [-0.05,0.39] | 0.69 | Anecdotal | Null | 45 |
| Accuracy | ASRM Mania | -0.07 | [-0.27,0.17] | 0.37 | Anecdotal | Null | 50 |
| Accuracy | BDI Depression | -0.09 | [-0.27,0.07] | 0.37 | Anecdotal | Null | 90 |
| Accuracy | SANS Amotivation | -0.04 | [-0.34,0.23] | 0.43 | Anecdotal | Null | 27 |
| Accuracy | SAPS Delusion | 0.08 | [-0.2,0.36] | 0.45 | Anecdotal | Null | 28 |
| Accuracy | SAPS Hallucination | -0.04 | [-0.32,0.24] | 0.43 | Anecdotal | Null | 28 |
| RT | BACS | -0.18 | [-0.4,0.04] | 0.79 | Anecdotal | Null | 46 |
| RT | MSCEIT | -0.23 | [-0.45,-0.02]\* | 1.40 | Anecdotal | Alternative | 45 |
| RT | ASRM Mania | 0.12 | [-0.1,0.34] | 0.48 | Anecdotal | Null | 50 |
| RT | BDI Depression | 0.09 | [-0.07,0.28] | 0.37 | Anecdotal | Null | 90 |
| RT | SANS Amotivation | -0.14 | [-0.43,0.13] | 0.58 | Anecdotal | Null | 27 |
| RT | SAPS Delusion | -0.17 | [-0.44,0.09] | 0.70 | Anecdotal | Null | 28 |
| RT | SAPS Hallucination | -0.18 | [-0.45,0.09] | 0.75 | Anecdotal | Null | 28 |
| SDT-Discriminability | BACS | 0.58 | [0.43,0.74]\* | 10708.55 | Extreme | Alternative | 46 |
| SDT-Discriminability | MSCEIT | 0.33 | [0.13,0.54]\* | 6.19 | Moderate | Alternative | 45 |
| SDT-Discriminability | ASRM Mania | -0.16 | [-0.37,0.04] | 0.66 | Anecdotal | Null | 50 |
| SDT-Discriminability | BDI Depression | -0.18 | [-0.34,-0.01]\* | 1.07 | Anecdotal | Alternative | 90 |
| SDT-Discriminability | SANS Amotivation | 0.03 | [-0.26,0.31] | 0.42 | Anecdotal | Null | 27 |
| SDT-Discriminability | SAPS Delusion | 0.14 | [-0.15,0.41] | 0.56 | Anecdotal | Null | 28 |
| SDT-Discriminability | SAPS Hallucination | 0.08 | [-0.21,0.35] | 0.46 | Anecdotal | Null | 28 |
| SDT-Criterion | BACS | -0.07 | [-0.29,0.15] | 0.37 | Anecdotal | Null | 46 |
| SDT-Criterion | MSCEIT | 0.10 | [-0.13,0.32] | 0.44 | Anecdotal | Null | 45 |
| SDT-Criterion | ASRM Mania | -0.16 | [-0.37,0.06] | 0.67 | Anecdotal | Null | 50 |
| SDT-Criterion | BDI Depression | -0.08 | [-0.24,0.09] | 0.32 | Moderate | Null | 90 |
| SDT-Criterion | SANS Amotivation | 0.18 | [-0.09,0.46] | 0.74 | Anecdotal | Null | 27 |
| SDT-Criterion | SAPS Delusion | 0.18 | [-0.1,0.45] | 0.71 | Anecdotal | Null | 28 |
| SDT-Criterion | SAPS Hallucination | 0.35 | [0.12,0.6]\* | 3.97 | Moderate | Alternative | 28 |
|  |  |  |  |  |  |  |  |
| --- | --- | --- | --- | --- | --- | --- | --- |
| Note: |  |  |  |  |  |  |  |
| \* = Credible association (i.e., 90% HDI does not contain zero); Mean = mean posterior estimate of correlation coefficient (equivalent to Pearson R); HDI = 90% HDI of the correlation coefficient; BF = Bayes Factor; BF\_Evidence\_Strength = Bayes Factor interpretation scheme based on Lee and Wagenmakers (2014); BF\_Evidence\_Favors = whether evidence favors the null hypothesis (i.e., the association between variable 1 and 2 is 0) or the alternative hypothesis (i.e., an association between variable 1 and 2 is not 0); N = Number of subjects with complete data included in correlation. |  |  |  |  |  |  |  |

#### 19.2 Sensitivity Analyses

Because some of the clinical metrics (SAPS, SANS) were zero-inflated
and clinical metrics and/or performance metrics contained potentially
influential observations, we ran sensitivity analyses to see if credible
relationships identified above held when 1) potentially influential
cases were removed, and 2) subjects with ‘0s’ for those
Hallucination/Delusion symptom dimensions were removed.

Results of sensitivity analyses show that the correlation between
hallucinations and drift bias for forward heads remains credible when
outliers and subjects with hallucinations scores = 0 are removed. In
fact, the correlation is strengthened somewhat. This suggests the
correlation is not merely the result of influential cases. However, the
correlations between hallucinations and drift bias for deviated heads–as
well as SDT-Criterion–were no longer credible after removing potentially
influential observations. As such, we will only interpret the
correlation between hallucinations and drift bias (forward). Results
also show that the correlation between delusions and start point is
stronger when potentially influential cases and subject with delusions =
‘0’ are removed, suggesting the relationship is not the by-product of
influential observations.

Correlations Between DDM Parameters, Traditional Metrics, and Clinical
Metrics After Removing Potentially Influential and Zero-Inflated Cases

| Sensitivity\_Analysis | Variable1 | Variable2 | Mean | HDI | BF | BF\_Evidence\_Strength | BF\_Evidence\_Favors | N |
| --- | --- | --- | --- | --- | --- | --- | --- | --- |
| Halluc\*Drift Bias (Forward) - Influential Cases Removed | SAPS Hallucination | Drift Bias (Forward) | -0.48 | [-0.69,-0.25]\* | 27.81 | Strong | Alternative | 27 |
| Halluc\*Drift Bias (Deviated) - Influential Cases Removed | SAPS Hallucination | Drift Bias (Deviated) | -0.13 | [-0.4,0.16] | 0.55 | Anecdotal | Null | 27 |
| Halluc\*SDT-Criterion - Influential Cases Removed | SAPS Hallucination | SDT-Criterion | 0.23 | [-0.07,0.48] | 1.07 | Anecdotal | Alternative | 27 |
| Halluc\*Drift Bias (Forward) - SAPS Halluc=0 Cases Removed | SAPS Hallucination | Drift Bias (Forward) | -0.41 | [-0.7,-0.11]\* | 3.83 | Moderate | Alternative | 17 |
| Delusion\*Start Point - Influential Cases Removed | SAPS Delusion | Start Point | 0.40 | [0.15,0.65]\* | 7.27 | Moderate | Alternative | 26 |
| Delusion\*Start Point - SAPS Delusion=0 Cases Removed | SAPS Delusion | Start Point | 0.33 | [0.03,0.64]\* | 2.44 | Anecdotal | Alternative | 21 |
| Amotiv\*Drift Bias (Forward) - Influential Cases Removed | SANS Amotivation | Drift Bias (Forward) | -0.22 | [-0.49,0.07] | 1.06 | Anecdotal | Alternative | 26 |
|  |  |  |  |  |  |  |  |  |
| --- | --- | --- | --- | --- | --- | --- | --- | --- |
| Note: |  |  |  |  |  |  |  |  |
| \* = Credible association (i.e., 90% HDI does not contain zero); Mean = mean posterior estimate of correlation coefficient (equivalent to Pearson R); HDI = 90% HDI of the correlation coefficient; BF = Bayes Factor; BF\_Evidence\_Strength = Bayes Factor interpretation scheme based on Lee and Wagenmakers (2014); N = Number of subjects with complete data included in correlation. |  |  |  |  |  |  |  |  |

#### 19.3 Post-Hoc Correlations

Following the primary correlations above, we performed a series of
post-hoc correlation analyses.

##### 19.3.1 Within SZ Only

We repeated the same key correlations of interest (from above) within
the SZ group only. We exclude SAPS hallucinations and delusions below
because the correlations above were already within SZ only. Credible
associations between drift rates and BACS and drift rate and
SDT-discriminability remain credible in SZ. The same is true for the
association between drift rate for forward heads and MSCEIT. Several
other credible associations also emerge but they are supported by only
anecdotal evidence and we chose not to interpret those.

Correlations (Within SZ Only) Between DDM Parameters, Traditional
Metrics, Clinical Metrics

| Variable1 | Variable2 | Mean | HDI | BF | BF\_Evidence\_Strength | BF\_Evidence\_Favors | N |
| --- | --- | --- | --- | --- | --- | --- | --- |
| Drift Rate (Forward) | BACS | 0.43 | [0.2,0.66]\* | 14.14 | Strong | Alternative | 28 |
| Drift Rate (Forward) | MSCEIT | 0.30 | [0.05,0.58]\* | 1.96 | Anecdotal | Alternative | 27 |
| Drift Rate (Forward) | BDI Depression | 0.07 | [-0.21,0.34] | 0.45 | Anecdotal | Null | 27 |
| Drift Rate (Deviated) | BACS | 0.36 | [0.12,0.61]\* | 4.19 | Moderate | Alternative | 28 |
| Drift Rate (Deviated) | MSCEIT | 0.17 | [-0.11,0.45] | 0.68 | Anecdotal | Null | 27 |
| Drift Rate (Deviated) | BDI Depression | -0.08 | [-0.37,0.18] | 0.48 | Anecdotal | Null | 27 |
| Threshold Separation | BACS | -0.10 | [-0.37,0.18] | 0.50 | Anecdotal | Null | 28 |
| Threshold Separation | MSCEIT | -0.03 | [-0.34,0.23] | 0.42 | Anecdotal | Null | 27 |
| Threshold Separation | BDI Depression | -0.15 | [-0.43,0.13] | 0.62 | Anecdotal | Null | 27 |
| Start Point | BACS | 0.25 | [0,0.53] | 1.19 | Anecdotal | Alternative | 28 |
| Start Point | MSCEIT | 0.24 | [-0.03,0.49] | 1.19 | Anecdotal | Alternative | 27 |
| Start Point | BDI Depression | 0.29 | [0.04,0.57]\* | 1.92 | Anecdotal | Alternative | 27 |
| Drift Bias (Forward) | BACS | -0.05 | [-0.34,0.22] | 0.43 | Anecdotal | Null | 28 |
| Drift Bias (Forward) | MSCEIT | -0.29 | [-0.55,-0.03]\* | 1.86 | Anecdotal | Alternative | 27 |
| Drift Bias (Forward) | BDI Depression | -0.20 | [-0.48,0.07] | 0.85 | Anecdotal | Null | 27 |
| Drift Bias (Deviated) | BACS | -0.03 | [-0.29,0.26] | 0.42 | Anecdotal | Null | 28 |
| Drift Bias (Deviated) | MSCEIT | -0.33 | [-0.58,-0.08]\* | 2.95 | Anecdotal | Alternative | 27 |
| Drift Bias (Deviated) | BDI Depression | -0.30 | [-0.55,-0.04]\* | 2.08 | Anecdotal | Alternative | 27 |
| Accuracy | BACS | 0.31 | [0.07,0.58]\* | 2.48 | Anecdotal | Alternative | 28 |
| Accuracy | MSCEIT | 0.09 | [-0.18,0.37] | 0.49 | Anecdotal | Null | 27 |
| Accuracy | BDI Depression | -0.06 | [-0.32,0.25] | 0.44 | Anecdotal | Null | 27 |
| RT | BACS | -0.08 | [-0.34,0.21] | 0.46 | Anecdotal | Null | 28 |
| RT | MSCEIT | -0.16 | [-0.45,0.11] | 0.66 | Anecdotal | Null | 27 |
| RT | BDI Depression | -0.14 | [-0.42,0.15] | 0.60 | Anecdotal | Null | 27 |
| SDT-Discriminability | BACS | 0.37 | [0.15,0.63]\* | 5.14 | Moderate | Alternative | 28 |
| SDT-Discriminability | MSCEIT | 0.25 | [-0.01,0.53] | 1.25 | Anecdotal | Alternative | 27 |
| SDT-Discriminability | BDI Depression | -0.02 | [-0.31,0.27] | 0.42 | Anecdotal | Null | 27 |
| SDT-Criterion | BACS | -0.02 | [-0.31,0.24] | 0.41 | Anecdotal | Null | 28 |
| SDT-Criterion | MSCEIT | 0.29 | [0.03,0.55]\* | 1.87 | Anecdotal | Alternative | 27 |
| SDT-Criterion | BDI Depression | 0.24 | [-0.03,0.51] | 1.11 | Anecdotal | Alternative | 27 |
|  |  |  |  |  |  |  |  |
| --- | --- | --- | --- | --- | --- | --- | --- |
| Note: |  |  |  |  |  |  |  |
| Mean = mean posterior estimate of correlation coefficient (equivalent to Pearson R); HDI = 90% HDI of the correlation coefficient; BF = Bayes Factor; BF\_Evidence\_Strength = Bayes Factor interpretation scheme based on Lee and Wagenmakers (2014); N = Number of subjects with complete data included in correlation. |  |  |  |  |  |  |  |

##### 19.3.2 Within BD Only

We repeated the same key correlations of interest (from above) within
the BD group only. We only include ASRM and BDI because the other
measures were not collected in BD. Results showed that neither ASRM or
BDI showed credible associations with DDM parameters or traditional
metrics in the BD sample.

Correlations (Within BD Only) Between DDM Parameters, Traditional
Metrics, Clinical Metrics

| Variable1 | Variable2 | Mean | HDI | BF | BF\_Evidence\_Strength | BF\_Evidence\_Favors | N |
| --- | --- | --- | --- | --- | --- | --- | --- |
| Drift Rate (Forward) | ASRM Mania | -0.07 | [-0.31,0.18] | 0.41 | Anecdotal | Null | 36 |
| Drift Rate (Forward) | BDI Depression | 0.07 | [-0.18,0.31] | 0.42 | Anecdotal | Null | 36 |
| Drift Rate (Deviated) | ASRM Mania | 0.05 | [-0.2,0.3] | 0.39 | Anecdotal | Null | 36 |
| Drift Rate (Deviated) | BDI Depression | 0.01 | [-0.22,0.28] | 0.37 | Anecdotal | Null | 36 |
| Threshold Separation | ASRM Mania | -0.04 | [-0.27,0.23] | 0.38 | Anecdotal | Null | 36 |
| Threshold Separation | BDI Depression | 0.11 | [-0.12,0.37] | 0.48 | Anecdotal | Null | 36 |
| Start Point | ASRM Mania | 0.06 | [-0.2,0.31] | 0.39 | Anecdotal | Null | 36 |
| Start Point | BDI Depression | 0.07 | [-0.19,0.31] | 0.42 | Anecdotal | Null | 36 |
| Drift Bias (Forward) | ASRM Mania | 0.10 | [-0.14,0.35] | 0.46 | Anecdotal | Null | 36 |
| Drift Bias (Forward) | BDI Depression | 0.16 | [-0.1,0.41] | 0.63 | Anecdotal | Null | 36 |
| Drift Bias (Deviated) | ASRM Mania | -0.01 | [-0.25,0.25] | 0.37 | Anecdotal | Null | 36 |
| Drift Bias (Deviated) | BDI Depression | 0.10 | [-0.15,0.35] | 0.46 | Anecdotal | Null | 36 |
| Accuracy | ASRM Mania | 0.00 | [-0.28,0.23] | 0.37 | Anecdotal | Null | 36 |
| Accuracy | BDI Depression | 0.13 | [-0.13,0.38] | 0.53 | Anecdotal | Null | 36 |
| RT | ASRM Mania | 0.04 | [-0.22,0.28] | 0.38 | Anecdotal | Null | 36 |
| RT | BDI Depression | 0.06 | [-0.21,0.3] | 0.40 | Anecdotal | Null | 36 |
| SDT-Discriminability | ASRM Mania | -0.09 | [-0.33,0.17] | 0.43 | Anecdotal | Null | 36 |
| SDT-Discriminability | BDI Depression | 0.04 | [-0.2,0.33] | 0.38 | Anecdotal | Null | 36 |
| SDT-Criterion | ASRM Mania | -0.08 | [-0.31,0.19] | 0.42 | Anecdotal | Null | 36 |
| SDT-Criterion | BDI Depression | -0.13 | [-0.39,0.12] | 0.54 | Anecdotal | Null | 36 |
|  |  |  |  |  |  |  |  |
| --- | --- | --- | --- | --- | --- | --- | --- |
| Note: |  |  |  |  |  |  |  |
| Mean = mean posterior estimate of correlation coefficient (equivalent to Pearson R); HDI = 90% HDI of the correlation coefficient; BF = Bayes Factor; BF\_Evidence\_Strength = Bayes Factor interpretation scheme based on Lee and Wagenmakers (2014); N = Number of subjects with complete data included in correlation. |  |  |  |  |  |  |  |

##### 19.3.3 Start Point and Paranoia

Given the relationship we observed between start point and SAPS
Delusions, we ran a post hoc follow-up test to examine whether this
relationship was evident within paranoia symptoms specifically. To
achieve this, we calculated a paranoia factor using results of a factor
analysis on SAPS items from Peralta (1999). That study identified that
two items from the SAPS – delusions of persecution and delusions of
reference – loaded onto a ‘paranoia’ factor. In our data, we generated a
paranoia factor by scaling participants’ scores on these two items by
the factor loadings of Peralta (1999) and summing them. Then we ran a
correlation (using a Bayesian approach) on this paranoia factor and the
DDM start point parameter.

Results revealed a credible positive correlation, such that SZ
patients with more paranoia also had higher start points (i.e., greater
initial self-referential biases) on the gaze task.

Correlation Between Start Point and SAPS Paranoia Factor

| Variable1 | Variable2 | Mean | HDI | BF | BF\_Evidence\_Strength | BF\_Evidence\_Favors | N |
| --- | --- | --- | --- | --- | --- | --- | --- |
| Start Point | SAPS Paranoia Factor | 0.31 | [0.08,0.59]\* | 2.37 | Anecdotal | Alternative | 28 |
|  |  |  |  |  |  |  |  |
| --- | --- | --- | --- | --- | --- | --- | --- |
| Note: |  |  |  |  |  |  |  |
| Mean = mean posterior estimate of correlation coefficient (equivalent to Pearson R); HDI = 90% HDI of the correlation coefficient; BF = Bayes Factor; BF\_Evidence\_Strength = Bayes Factor interpretation scheme based on Lee and Wagenmakers (2014); N = Number of subjects with complete data included in correlation. |  |  |  |  |  |  |  |

##### 19.3.4 Antipsychotics and Performance

To assess whether antipsychotic doses (CPZeq) were related to
measures that tap processing speed, we ran correlations between CPZeq
and measures that were sensitive to processing speed. This was done
exclusively within participants in the SZ and BD groups taking
antipsychotic medicaitons. This included BACS, DDM parameters, and
traditional performance metrics on the gaze task. MSCEIT was not
included as it does not impose a time limit on participants.

Results did not show credible associations between CPZ dose and any
of the DDM parameters, BACS scores, or other gaze task performance
metrics. This suggested that observed results were not merely the result
of the influence of antipsychotic dosing.

Correlations Between Antipsychotic Dose and Performance

| Variable1 | Variable2 | Mean | HDI | BF | BF\_Evidence\_Strength | BF\_Evidence\_Favors | N |
| --- | --- | --- | --- | --- | --- | --- | --- |
| CPZeq | BACS | -0.03 | [-0.32,0.25] | 0.43 | Anecdotal | Null | 27 |
| CPZeq | Threshold\_Separation | 0.14 | [-0.05,0.35] | 0.58 | Anecdotal | Null | 60 |
| CPZeq | Start\_Point | -0.06 | [-0.27,0.13] | 0.33 | Moderate | Null | 60 |
| CPZeq | Drift\_Rate\_Forward | -0.14 | [-0.35,0.05] | 0.57 | Anecdotal | Null | 60 |
| CPZeq | Drift\_Rate\_Deviated | -0.07 | [-0.27,0.14] | 0.34 | Anecdotal | Null | 60 |
| CPZeq | Drift\_Bias\_Forward | -0.13 | [-0.33,0.06] | 0.54 | Anecdotal | Null | 60 |
| CPZeq | Drift\_Bias\_Deviated | 0.11 | [-0.09,0.32] | 0.44 | Anecdotal | Null | 60 |
| CPZeq | Accuracy | -0.13 | [-0.35,0.05] | 0.54 | Anecdotal | Null | 60 |
| CPZeq | RT | 0.09 | [-0.12,0.28] | 0.37 | Anecdotal | Null | 60 |
| CPZeq | Criterion | 0.01 | [-0.18,0.22] | 0.29 | Moderate | Null | 60 |
| CPZeq | Discriminability | -0.12 | [-0.32,0.08] | 0.51 | Anecdotal | Null | 60 |
|  |  |  |  |  |  |  |  |
| --- | --- | --- | --- | --- | --- | --- | --- |
| Note: |  |  |  |  |  |  |  |
| Mean = mean posterior estimate of correlation coefficient (equivalent to Pearson R); HDI = 90% HDI of the correlation coefficient; BF = Bayes Factor; BF\_Evidence\_Strength = Bayes Factor interpretation scheme based on Lee and Wagenmakers (2014); N = Number of subjects with complete data included in correlation. |  |  |  |  |  |  |  |

### 20 Regressions

We ran separate hierarchical linear regressions to assess whether any
of the DDM parameters and/or traditional gaze task performance metrics
(accuracy, RT, SDT Criterion, SDT discriminability) could predict social
functioning (Social Adjustment Scale-SR, Social/Leisure Sub scale)
across SZ and HC, above and beyond diagnosis and common measures of
general (BACS) and emotion-based social cognition (MSCEIT). We assessed
these predictors on whether: 1) They were credible predictors of social
functioning. Credible predictors were those in which the 90% HDI of the
predictor coefficient did not contain zero. 2) They improved the
out-of-sample predictive accuracy. This was done using LOO model
comparisons.

These regression models were run in Stan via brms (Burkner, 2017)
using standardized predictors and weakly informative priors (i.e.,
Normal(0,1) for each predictor). Models were sampled using 1000 warmup
samples and 4000 postwarmup draws for each of 4 chains, resulting in
16000 total post warmup samples. The same procedure for assessing
convergence (described above) indicated that parameters of all models
had converged to their target distributions.

#### 20.1 DDM Parameters Predicting Social Functioning

Start point predicts social functioning above and beyond diagnosis,
general cognition, and social cognition. The model including start point
showed increases in predictive accuracy relative to the null model, but
the strength of evidence for both the full model (relative to the null)
and strength of evidence for the start point predictor are both only
anecdotal.

Bayesian Regression: Predicting Social Functioning from DDM Parameters

| Model | Ref. Model | LOO ELPD | LOO ELPD SE | ΔELPD | ΔELPD SE | Model BF | Model BF Strength/ Direction | Predictor | Pred BF | Pred BF Strength/ Direction | Pred Mean [90% HDI] |
| --- | --- | --- | --- | --- | --- | --- | --- | --- | --- | --- | --- |
| 1 | 1 | -59.16 | 3.94 | – | – | – | –/– |  |  |  |  |
|  |  |  |  |  |  |  |  | Intercept | 0.13 | Null/Moderate | 0 [-0.21, 0.23] |
|  |  |  |  |  |  |  |  | SZ\_Dummy | 12.99 | Alternative/Strong | -0.5 [-0.77, -0.22]\* |
|  |  |  |  |  |  |  |  | BACS | 0.19 | Null/Moderate | 0.1 [-0.17, 0.37] |
|  |  |  |  |  |  |  |  | MSCEIT | 0.16 | Null/Moderate | -0.01 [-0.27, 0.25] |
| 2 | 1 | -59.66 | 4.08 | -0.5 | 0.85 | 0.27 | Null/Moderate |  |  |  |  |
|  |  |  |  |  |  |  |  | Intercept | 0.13 | Null/Moderate | 0 [-0.21, 0.22] |
|  |  |  |  |  |  |  |  | SZ\_Dummy | 15.06 | Alternative/Strong | -0.53 [-0.81, -0.24]\* |
|  |  |  |  |  |  |  |  | BACS | 0.31 | Null/Moderate | 0.18 [-0.12, 0.51] |
|  |  |  |  |  |  |  |  | MSCEIT | 0.16 | Null/Moderate | 0 [-0.28, 0.25] |
|  |  |  |  |  |  |  |  | Drift\_Rate\_Forward | 0.27 | Null/Moderate | -0.16 [-0.47, 0.15] |
| 3 | 1 | -59.12 | 3.73 | 0.03 | 1.15 | 0.36 | Null/Anecdotal |  |  |  |  |
|  |  |  |  |  |  |  |  | Intercept | 0.13 | Null/Moderate | 0 [-0.23, 0.21] |
|  |  |  |  |  |  |  |  | SZ\_Dummy | 21.49 | Alternative/Strong | -0.54 [-0.82, -0.27]\* |
|  |  |  |  |  |  |  |  | BACS | 0.38 | Null/Anecdotal | 0.22 [-0.1, 0.55] |
|  |  |  |  |  |  |  |  | MSCEIT | 0.17 | Null/Moderate | -0.05 [-0.31, 0.22] |
|  |  |  |  |  |  |  |  | Drift\_Rate\_Deviated | 0.36 | Null/Anecdotal | -0.21 [-0.51, 0.07] |
| 4 | 1 | -60.03 | 4.02 | -0.88 | 0.65 | 0.16 | Null/Moderate |  |  |  |  |
|  |  |  |  |  |  |  |  | Intercept | 0.13 | Null/Moderate | 0 [-0.21, 0.22] |
|  |  |  |  |  |  |  |  | SZ\_Dummy | 14.06 | Alternative/Strong | -0.51 [-0.77, -0.22]\* |
|  |  |  |  |  |  |  |  | BACS | 0.19 | Null/Moderate | 0.08 [-0.2, 0.36] |
|  |  |  |  |  |  |  |  | MSCEIT | 0.16 | Null/Moderate | 0 [-0.28, 0.27] |
|  |  |  |  |  |  |  |  | Drift\_Bias\_Forward | 0.16 | Null/Moderate | 0.08 [-0.14, 0.31] |
| 5 | 1 | -58.06 | 4.24 | 1.1 | 1.42 | 0.75 | Null/Anecdotal |  |  |  |  |
|  |  |  |  |  |  |  |  | Intercept | 0.13 | Null/Moderate | 0 [-0.22, 0.2] |
|  |  |  |  |  |  |  |  | SZ\_Dummy | 22.74 | Alternative/Strong | -0.54 [-0.83, -0.29]\* |
|  |  |  |  |  |  |  |  | BACS | 0.18 | Null/Moderate | 0.08 [-0.2, 0.34] |
|  |  |  |  |  |  |  |  | MSCEIT | 0.16 | Null/Moderate | 0.02 [-0.24, 0.28] |
|  |  |  |  |  |  |  |  | Drift\_Bias\_Deviated | 0.76 | Null/Anecdotal | 0.25 [0.03, 0.46]\* |
| 6 | 1 | -57.14 | 3.39 | 2.02 | 1.99 | 1.55 | Alternative/Anecdotal |  |  |  |  |
|  |  |  |  |  |  |  |  | Intercept | 0.12 | Null/Moderate | 0 [-0.2, 0.2] |
|  |  |  |  |  |  |  |  | SZ\_Dummy | 17.98 | Alternative/Strong | -0.51 [-0.78, -0.25]\* |
|  |  |  |  |  |  |  |  | BACS | 0.36 | Null/Anecdotal | 0.2 [-0.07, 0.47] |
|  |  |  |  |  |  |  |  | MSCEIT | 0.15 | Null/Moderate | -0.03 [-0.28, 0.22] |
|  |  |  |  |  |  |  |  | Start\_Point | 1.54 | Alternative/Anecdotal | -0.3 [-0.52, -0.07]\* |
| 7 | 1 | -60.26 | 3.9 | -1.1 | 0.26 | 0.14 | Null/Moderate |  |  |  |  |
|  |  |  |  |  |  |  |  | Intercept | 0.13 | Null/Moderate | 0 [-0.21, 0.22] |
|  |  |  |  |  |  |  |  | SZ\_Dummy | 9.62 | Alternative/Moderate | -0.5 [-0.79, -0.23]\* |
|  |  |  |  |  |  |  |  | BACS | 0.2 | Null/Moderate | 0.1 [-0.19, 0.36] |
|  |  |  |  |  |  |  |  | MSCEIT | 0.16 | Null/Moderate | -0.01 [-0.3, 0.25] |
|  |  |  |  |  |  |  |  | Threshold\_Separation | 0.13 | Null/Moderate | -0.01 [-0.24, 0.22] |

#### 20.2 Sensitivity Analysis (Control for CPZeq)

Results are the same when we control for antipsychotic dose.

Bayesian Regression (Sensitivity Analysis): Predicting Social
Functioning from DDM Parameters after Controlling for Antipsychotic Dose

| Model | Ref. Model | LOO ELPD | LOO ELPD SE | ΔELPD | ΔELPD SE | Model BF | Model BF Strength/ Direction | Predictor | Pred BF | Pred BF Strength/ Direction | Pred Mean [90% HDI] |
| --- | --- | --- | --- | --- | --- | --- | --- | --- | --- | --- | --- |
| 1 | 1 | -60.3 | 4.14 | – | – | – | –/– |  |  |  |  |
|  |  |  |  |  |  |  |  | Intercept | 0.13 | Null/Moderate | 0 [-0.22, 0.22] |
|  |  |  |  |  |  |  |  | SZ\_Dummy | 8.67 | Alternative/Moderate | -0.53 [-0.84, -0.23]\* |
|  |  |  |  |  |  |  |  | BACS | 0.2 | Null/Moderate | 0.1 [-0.18, 0.38] |
|  |  |  |  |  |  |  |  | MSCEIT | 0.16 | Null/Moderate | -0.01 [-0.29, 0.26] |
|  |  |  |  |  |  |  |  | CPZeq | 0.18 | Null/Moderate | 0.06 [-0.2, 0.34] |
| 2 | 1 | -61 | 4.15 | -0.7 | 0.73 | 0.27 | Null/Moderate |  |  |  |  |
|  |  |  |  |  |  |  |  | Intercept | 0.14 | Null/Moderate | 0 [-0.22, 0.22] |
|  |  |  |  |  |  |  |  | SZ\_Dummy | 8.79 | Alternative/Moderate | -0.54 [-0.84, -0.22]\* |
|  |  |  |  |  |  |  |  | BACS | 0.3 | Null/Moderate | 0.17 [-0.16, 0.5] |
|  |  |  |  |  |  |  |  | MSCEIT | 0.16 | Null/Moderate | 0 [-0.29, 0.27] |
|  |  |  |  |  |  |  |  | CPZeq | 0.18 | Null/Moderate | 0.01 [-0.28, 0.31] |
|  |  |  |  |  |  |  |  | Drift\_Rate\_Forward | 0.27 | Null/Moderate | -0.15 [-0.5, 0.19] |
| 3 | 1 | -60.44 | 3.83 | -0.14 | 1.1 | 0.35 | Null/Anecdotal |  |  |  |  |
|  |  |  |  |  |  |  |  | Intercept | 0.13 | Null/Moderate | 0 [-0.22, 0.22] |
|  |  |  |  |  |  |  |  | SZ\_Dummy | 10.79 | Alternative/Strong | -0.55 [-0.85, -0.24]\* |
|  |  |  |  |  |  |  |  | BACS | 0.37 | Null/Anecdotal | 0.22 [-0.11, 0.55] |
|  |  |  |  |  |  |  |  | MSCEIT | 0.18 | Null/Moderate | -0.04 [-0.32, 0.23] |
|  |  |  |  |  |  |  |  | CPZeq | 0.16 | Null/Moderate | 0.02 [-0.25, 0.3] |
|  |  |  |  |  |  |  |  | Drift\_Rate\_Deviated | 0.34 | Null/Anecdotal | -0.2 [-0.51, 0.09] |
| 4 | 1 | -61.23 | 4.34 | -0.93 | 0.74 | 0.17 | Null/Moderate |  |  |  |  |
|  |  |  |  |  |  |  |  | Intercept | 0.13 | Null/Moderate | 0 [-0.23, 0.21] |
|  |  |  |  |  |  |  |  | SZ\_Dummy | 11.49 | Alternative/Strong | -0.55 [-0.87, -0.24]\* |
|  |  |  |  |  |  |  |  | BACS | 0.19 | Null/Moderate | 0.08 [-0.19, 0.37] |
|  |  |  |  |  |  |  |  | MSCEIT | 0.16 | Null/Moderate | 0.01 [-0.27, 0.27] |
|  |  |  |  |  |  |  |  | CPZeq | 0.18 | Null/Moderate | 0.07 [-0.21, 0.34] |
|  |  |  |  |  |  |  |  | Drift\_Bias\_Forward | 0.16 | Null/Moderate | 0.09 [-0.15, 0.31] |
| 5 | 1 | -59.53 | 4.42 | 0.77 | 1.55 | 0.69 | Null/Anecdotal |  |  |  |  |
|  |  |  |  |  |  |  |  | Intercept | 0.13 | Null/Moderate | 0 [-0.21, 0.22] |
|  |  |  |  |  |  |  |  | SZ\_Dummy | 14.38 | Alternative/Strong | -0.56 [-0.86, -0.25]\* |
|  |  |  |  |  |  |  |  | BACS | 0.18 | Null/Moderate | 0.07 [-0.18, 0.35] |
|  |  |  |  |  |  |  |  | MSCEIT | 0.16 | Null/Moderate | 0.02 [-0.25, 0.28] |
|  |  |  |  |  |  |  |  | CPZeq | 0.16 | Null/Moderate | 0.03 [-0.24, 0.29] |
|  |  |  |  |  |  |  |  | Drift\_Bias\_Deviated | 0.74 | Null/Anecdotal | 0.24 [0.01, 0.46]\* |
| 6 | 1 | -58.85 | 3.66 | 1.45 | 2.07 | 1.41 | Alternative/Anecdotal |  |  |  |  |
|  |  |  |  |  |  |  |  | Intercept | 0.13 | Null/Moderate | 0 [-0.21, 0.22] |
|  |  |  |  |  |  |  |  | SZ\_Dummy | 10.92 | Alternative/Strong | -0.53 [-0.82, -0.23]\* |
|  |  |  |  |  |  |  |  | BACS | 0.36 | Null/Anecdotal | 0.21 [-0.06, 0.5] |
|  |  |  |  |  |  |  |  | MSCEIT | 0.16 | Null/Moderate | -0.02 [-0.28, 0.23] |
|  |  |  |  |  |  |  |  | CPZeq | 0.16 | Null/Moderate | 0.03 [-0.22, 0.31] |
|  |  |  |  |  |  |  |  | Start\_Point | 1.35 | Alternative/Anecdotal | -0.3 [-0.53, -0.07]\* |
| 7 | 1 | -61.42 | 4.14 | -1.12 | 0.27 | 0.14 | Null/Moderate |  |  |  |  |
|  |  |  |  |  |  |  |  | Intercept | 0.14 | Null/Moderate | 0 [-0.22, 0.22] |
|  |  |  |  |  |  |  |  | SZ\_Dummy | 9.36 | Alternative/Moderate | -0.53 [-0.84, -0.21]\* |
|  |  |  |  |  |  |  |  | BACS | 0.2 | Null/Moderate | 0.1 [-0.19, 0.37] |
|  |  |  |  |  |  |  |  | MSCEIT | 0.16 | Null/Moderate | 0 [-0.27, 0.27] |
|  |  |  |  |  |  |  |  | CPZeq | 0.18 | Null/Moderate | 0.06 [-0.22, 0.33] |
|  |  |  |  |  |  |  |  | Threshold\_Separation | 0.14 | Null/Moderate | -0.01 [-0.24, 0.21] |

#### 20.3 Post-Hoc Analyses

Results look similar in SZ. The direction of the relationship between
the start point predictor and outcome is similar, but no longer
credible. This isn’t surprising given the loss of small sample size from
doing this within groups.

##### 20.3.1 DDM Parameters as Predictors (SZ Only)

Bayesian Regression: Predicting Social Functioning from DDM Parameters
(Within SZ Only)

| Model | Ref. Model | LOO ELPD | LOO ELPD SE | ΔELPD | ΔELPD SE | Model BF | Model BF Strength/ Direction | Predictor | Pred BF | Pred BF Strength/ Direction | Pred Mean [90% HDI] |
| --- | --- | --- | --- | --- | --- | --- | --- | --- | --- | --- | --- |
| 1 | 1 | -38.23 | 3.74 | – | – | – | –/– |  |  |  |  |
|  |  |  |  |  |  |  |  | Intercept | 0.17 | Null/Moderate | 0 [-0.31, 0.31] |
|  |  |  |  |  |  |  |  | BACS | 0.72 | Null/Anecdotal | 0.32 [-0.03, 0.65] |
|  |  |  |  |  |  |  |  | MSCEIT | 1.46 | Alternative/Anecdotal | -0.41 [-0.76, -0.08]\* |
| 2 | 1 | -39.22 | 3.9 | -0.99 | 0.73 | 0.27 | Null/Moderate |  |  |  |  |
|  |  |  |  |  |  |  |  | Intercept | 0.19 | Null/Moderate | 0 [-0.33, 0.31] |
|  |  |  |  |  |  |  |  | BACS | 0.9 | Null/Anecdotal | 0.38 [0, 0.77] |
|  |  |  |  |  |  |  |  | MSCEIT | 1.24 | Alternative/Anecdotal | -0.39 [-0.75, -0.04]\* |
|  |  |  |  |  |  |  |  | Drift\_Rate\_Forward | 0.27 | Null/Moderate | -0.13 [-0.51, 0.25] |
| 3 | 1 | -38.67 | 3.66 | -0.44 | 0.86 | 0.33 | Null/Anecdotal |  |  |  |  |
|  |  |  |  |  |  |  |  | Intercept | 0.19 | Null/Moderate | 0 [-0.31, 0.32] |
|  |  |  |  |  |  |  |  | BACS | 1.21 | Alternative/Anecdotal | 0.41 [0.03, 0.8]\* |
|  |  |  |  |  |  |  |  | MSCEIT | 1.47 | Alternative/Anecdotal | -0.41 [-0.76, -0.05]\* |
|  |  |  |  |  |  |  |  | Drift\_Rate\_Deviated | 0.33 | Null/Anecdotal | -0.19 [-0.55, 0.16] |
| 4 | 1 | -39.48 | 3.71 | -1.25 | 0.52 | 0.21 | Null/Moderate |  |  |  |  |
|  |  |  |  |  |  |  |  | Intercept | 0.19 | Null/Moderate | 0 [-0.33, 0.3] |
|  |  |  |  |  |  |  |  | BACS | 0.7 | Null/Anecdotal | 0.32 [-0.04, 0.67] |
|  |  |  |  |  |  |  |  | MSCEIT | 1.22 | Alternative/Anecdotal | -0.42 [-0.79, -0.03]\* |
|  |  |  |  |  |  |  |  | Drift\_Bias\_Forward | 0.21 | Null/Moderate | -0.02 [-0.36, 0.33] |
| 5 | 1 | -39.18 | 3.61 | -0.95 | 0.27 | 0.21 | Null/Moderate |  |  |  |  |
|  |  |  |  |  |  |  |  | Intercept | 0.19 | Null/Moderate | 0 [-0.32, 0.32] |
|  |  |  |  |  |  |  |  | BACS | 0.68 | Null/Anecdotal | 0.32 [-0.04, 0.68] |
|  |  |  |  |  |  |  |  | MSCEIT | 1.08 | Alternative/Anecdotal | -0.41 [-0.79, -0.02]\* |
|  |  |  |  |  |  |  |  | Drift\_Bias\_Deviated | 0.22 | Null/Moderate | 0.01 [-0.35, 0.36] |
| 6 | 1 | -38.06 | 3.23 | 0.16 | 1.27 | 0.57 | Null/Anecdotal |  |  |  |  |
|  |  |  |  |  |  |  |  | Intercept | 0.18 | Null/Moderate | 0 [-0.31, 0.31] |
|  |  |  |  |  |  |  |  | BACS | 1.07 | Alternative/Anecdotal | 0.38 [0.03, 0.73]\* |
|  |  |  |  |  |  |  |  | MSCEIT | 0.86 | Null/Anecdotal | -0.35 [-0.69, 0] |
|  |  |  |  |  |  |  |  | Start\_Point | 0.58 | Null/Anecdotal | -0.29 [-0.63, 0.05] |
| 7 | 1 | -38.92 | 3.52 | -0.69 | 0.48 | 0.22 | Null/Moderate |  |  |  |  |
|  |  |  |  |  |  |  |  | Intercept | 0.19 | Null/Moderate | 0 [-0.3, 0.32] |
|  |  |  |  |  |  |  |  | BACS | 0.75 | Null/Anecdotal | 0.33 [-0.02, 0.69] |
|  |  |  |  |  |  |  |  | MSCEIT | 1.5 | Alternative/Anecdotal | -0.42 [-0.77, -0.06]\* |
|  |  |  |  |  |  |  |  | Threshold\_Separation | 0.22 | Null/Moderate | 0.09 [-0.23, 0.42] |

##### 20.3.2 DDM Parameters as Predictors (HC Only)

Bayesian Regression: Predicting Social Functioning from DDM Parameters
(Within HC Only)

| Model | Ref. Model | LOO ELPD | LOO ELPD SE | ΔELPD | ΔELPD SE | Model BF | Model BF Strength/ Direction | Predictor | Pred BF | Pred BF Strength/ Direction | Pred Mean [90% HDI] |
| --- | --- | --- | --- | --- | --- | --- | --- | --- | --- | --- | --- |
| 1 | 1 | -26.22 | 2.11 | – | – | – | –/– |  |  |  |  |
|  |  |  |  |  |  |  |  | Intercept | 0.22 | Null/Moderate | 0 [-0.39, 0.35] |
|  |  |  |  |  |  |  |  | BACS | 0.24 | Null/Moderate | -0.09 [-0.47, 0.3] |
|  |  |  |  |  |  |  |  | MSCEIT | 2.19 | Alternative/Anecdotal | 0.48 [0.1, 0.86]\* |
| 2 | 1 | -27.54 | 2.43 | -1.33 | 1.38 | 0.34 | Null/Anecdotal |  |  |  |  |
|  |  |  |  |  |  |  |  | Intercept | 0.23 | Null/Moderate | 0 [-0.39, 0.37] |
|  |  |  |  |  |  |  |  | BACS | 0.36 | Null/Anecdotal | -0.17 [-0.67, 0.33] |
|  |  |  |  |  |  |  |  | MSCEIT | 2.07 | Alternative/Anecdotal | 0.5 [0.1, 0.91]\* |
|  |  |  |  |  |  |  |  | Drift\_Rate\_Forward | 0.35 | Null/Anecdotal | 0.14 [-0.36, 0.65] |
| 3 | 1 | -27.02 | 2.21 | -0.8 | 0.32 | 0.4 | Null/Anecdotal |  |  |  |  |
|  |  |  |  |  |  |  |  | Intercept | 0.22 | Null/Moderate | 0 [-0.39, 0.36] |
|  |  |  |  |  |  |  |  | BACS | 0.42 | Null/Anecdotal | -0.19 [-0.76, 0.4] |
|  |  |  |  |  |  |  |  | MSCEIT | 2.04 | Alternative/Anecdotal | 0.52 [0.09, 0.94]\* |
|  |  |  |  |  |  |  |  | Drift\_Rate\_Deviated | 0.39 | Null/Anecdotal | 0.15 [-0.46, 0.74] |
| 4 | 1 | -27.29 | 1.97 | -1.08 | 0.39 | 0.26 | Null/Moderate |  |  |  |  |
|  |  |  |  |  |  |  |  | Intercept | 0.23 | Null/Moderate | 0 [-0.38, 0.4] |
|  |  |  |  |  |  |  |  | BACS | 0.27 | Null/Moderate | -0.08 [-0.52, 0.35] |
|  |  |  |  |  |  |  |  | MSCEIT | 1.63 | Alternative/Anecdotal | 0.48 [0.05, 0.88]\* |
|  |  |  |  |  |  |  |  | Drift\_Bias\_Forward | 0.28 | Null/Moderate | -0.02 [-0.46, 0.44] |
| 5 | 1 | -25.81 | 2.15 | 0.41 | 0.83 | 0.85 | Null/Anecdotal |  |  |  |  |
|  |  |  |  |  |  |  |  | Intercept | 0.22 | Null/Moderate | 0 [-0.37, 0.34] |
|  |  |  |  |  |  |  |  | BACS | 0.27 | Null/Moderate | -0.14 [-0.51, 0.24] |
|  |  |  |  |  |  |  |  | MSCEIT | 0.69 | Null/Anecdotal | 0.34 [-0.06, 0.73] |
|  |  |  |  |  |  |  |  | Drift\_Bias\_Deviated | 0.85 | Null/Anecdotal | 0.37 [-0.03, 0.76] |
| 6 | 1 | -27 | 1.92 | -0.78 | 0.29 | 0.29 | Null/Moderate |  |  |  |  |
|  |  |  |  |  |  |  |  | Intercept | 0.23 | Null/Moderate | 0 [-0.38, 0.38] |
|  |  |  |  |  |  |  |  | BACS | 0.25 | Null/Moderate | -0.06 [-0.48, 0.35] |
|  |  |  |  |  |  |  |  | MSCEIT | 1.09 | Alternative/Anecdotal | 0.44 [-0.01, 0.88] |
|  |  |  |  |  |  |  |  | Start\_Point | 0.28 | Null/Moderate | -0.07 [-0.54, 0.39] |
| 7 | 1 | -27.52 | 2.18 | -1.31 | 0.74 | 0.27 | Null/Moderate |  |  |  |  |
|  |  |  |  |  |  |  |  | Intercept | 0.23 | Null/Moderate | 0 [-0.4, 0.38] |
|  |  |  |  |  |  |  |  | BACS | 0.24 | Null/Moderate | -0.06 [-0.46, 0.34] |
|  |  |  |  |  |  |  |  | MSCEIT | 1.84 | Alternative/Anecdotal | 0.47 [0.09, 0.87]\* |
|  |  |  |  |  |  |  |  | Threshold\_Separation | 0.29 | Null/Moderate | -0.11 [-0.52, 0.28] |

##### 20.3.3 Traditional Metrics as Predictors (Full Sample)

None of the traditional metrics can predict social functioning above
and beyond diagnosis, BACS, and MSCEIT.

Bayesian Regression: Predicting Social Functioning from Traditional
Metrics (Full Sample)

| Model | Ref. Model | LOO ELPD | LOO ELPD SE | ΔELPD | ΔELPD SE | Model BF | Model BF Strength/ Direction | Predictor | Pred BF | Pred BF Strength/ Direction | Pred Mean [90% HDI] |
| --- | --- | --- | --- | --- | --- | --- | --- | --- | --- | --- | --- |
| 1 | 1 | -59.16 | 3.94 | – | – | – | –/– |  |  |  |  |
|  |  |  |  |  |  |  |  | Intercept | 0.13 | Null/Moderate | 0 [-0.21, 0.23] |
|  |  |  |  |  |  |  |  | SZ\_Dummy | 12.99 | Alternative/Strong | -0.5 [-0.77, -0.22]\* |
|  |  |  |  |  |  |  |  | BACS | 0.19 | Null/Moderate | 0.1 [-0.17, 0.37] |
|  |  |  |  |  |  |  |  | MSCEIT | 0.16 | Null/Moderate | -0.01 [-0.27, 0.25] |
| 2 | 1 | -59.87 | 3.86 | -0.72 | 0.38 | 0.19 | Null/Moderate |  |  |  |  |
|  |  |  |  |  |  |  |  | Intercept | 0.13 | Null/Moderate | 0 [-0.22, 0.22] |
|  |  |  |  |  |  |  |  | SZ\_Dummy | 13.41 | Alternative/Strong | -0.5 [-0.77, -0.21]\* |
|  |  |  |  |  |  |  |  | BACS | 0.27 | Null/Moderate | 0.15 [-0.17, 0.48] |
|  |  |  |  |  |  |  |  | MSCEIT | 0.16 | Null/Moderate | -0.02 [-0.3, 0.25] |
|  |  |  |  |  |  |  |  | Accuracy | 0.18 | Null/Moderate | -0.09 [-0.37, 0.18] |
| 3 | 1 | -60.33 | 4.47 | -1.18 | 1.4 | 0.18 | Null/Moderate |  |  |  |  |
|  |  |  |  |  |  |  |  | Intercept | 0.13 | Null/Moderate | 0 [-0.22, 0.22] |
|  |  |  |  |  |  |  |  | SZ\_Dummy | 12.68 | Alternative/Strong | -0.53 [-0.82, -0.25]\* |
|  |  |  |  |  |  |  |  | BACS | 0.19 | Null/Moderate | 0.09 [-0.18, 0.37] |
|  |  |  |  |  |  |  |  | MSCEIT | 0.16 | Null/Moderate | 0 [-0.27, 0.26] |
|  |  |  |  |  |  |  |  | RT | 0.17 | Null/Moderate | 0.09 [-0.13, 0.33] |
| 4 | 1 | -59.58 | 4.34 | -0.42 | 1.03 | 0.24 | Null/Moderate |  |  |  |  |
|  |  |  |  |  |  |  |  | Intercept | 0.13 | Null/Moderate | 0 [-0.22, 0.21] |
|  |  |  |  |  |  |  |  | SZ\_Dummy | 15.47 | Alternative/Strong | -0.51 [-0.78, -0.23]\* |
|  |  |  |  |  |  |  |  | BACS | 0.18 | Null/Moderate | 0.06 [-0.21, 0.34] |
|  |  |  |  |  |  |  |  | MSCEIT | 0.16 | Null/Moderate | 0.01 [-0.25, 0.28] |
|  |  |  |  |  |  |  |  | Criterion | 0.23 | Null/Moderate | -0.14 [-0.36, 0.08] |
| 5 | 1 | -59.65 | 4.2 | -0.49 | 0.8 | 0.28 | Null/Moderate |  |  |  |  |
|  |  |  |  |  |  |  |  | Intercept | 0.13 | Null/Moderate | 0 [-0.22, 0.22] |
|  |  |  |  |  |  |  |  | SZ\_Dummy | 11.57 | Alternative/Strong | -0.51 [-0.79, -0.24]\* |
|  |  |  |  |  |  |  |  | BACS | 0.32 | Null/Moderate | 0.2 [-0.12, 0.53] |
|  |  |  |  |  |  |  |  | MSCEIT | 0.17 | Null/Moderate | -0.01 [-0.28, 0.26] |
|  |  |  |  |  |  |  |  | Discriminability | 0.28 | Null/Moderate | -0.16 [-0.46, 0.13] |

##### 20.3.4 Traditional Metrics as Predictors (SZ Only)

Bayesian Regression: Predicting Social Functioning from Traditional
Metrics (Within SZ Only)

| Model | Ref. Model | LOO ELPD | LOO ELPD SE | ΔELPD | ΔELPD SE | Model BF | Model BF Strength/ Direction | Predictor | Pred BF | Pred BF Strength/ Direction | Pred Mean [90% HDI] |
| --- | --- | --- | --- | --- | --- | --- | --- | --- | --- | --- | --- |
| 1 | 1 | -38.23 | 3.74 | – | – | – | –/– |  |  |  |  |
|  |  |  |  |  |  |  |  | Intercept | 0.17 | Null/Moderate | 0 [-0.31, 0.31] |
|  |  |  |  |  |  |  |  | BACS | 0.72 | Null/Anecdotal | 0.32 [-0.03, 0.65] |
|  |  |  |  |  |  |  |  | MSCEIT | 1.46 | Alternative/Anecdotal | -0.41 [-0.76, -0.08]\* |
| 2 | 1 | -38.82 | 3.89 | -0.59 | 0.54 | 0.29 | Null/Moderate |  |  |  |  |
|  |  |  |  |  |  |  |  | Intercept | 0.19 | Null/Moderate | 0 [-0.31, 0.31] |
|  |  |  |  |  |  |  |  | BACS | 1.01 | Alternative/Anecdotal | 0.39 [0.02, 0.78]\* |
|  |  |  |  |  |  |  |  | MSCEIT | 1.42 | Alternative/Anecdotal | -0.42 [-0.78, -0.08]\* |
|  |  |  |  |  |  |  |  | Accuracy | 0.29 | Null/Moderate | -0.17 [-0.51, 0.19] |
| 3 | 1 | -39.76 | 4.08 | -1.53 | 1.04 | 0.21 | Null/Moderate |  |  |  |  |
|  |  |  |  |  |  |  |  | Intercept | 0.2 | Null/Moderate | 0 [-0.32, 0.31] |
|  |  |  |  |  |  |  |  | BACS | 0.66 | Null/Anecdotal | 0.32 [-0.03, 0.67] |
|  |  |  |  |  |  |  |  | MSCEIT | 1.28 | Alternative/Anecdotal | -0.4 [-0.76, -0.05]\* |
|  |  |  |  |  |  |  |  | RT | 0.21 | Null/Moderate | 0.07 [-0.27, 0.38] |
| 4 | 1 | -39.27 | 3.59 | -1.05 | 0.67 | 0.23 | Null/Moderate |  |  |  |  |
|  |  |  |  |  |  |  |  | Intercept | 0.19 | Null/Moderate | 0 [-0.32, 0.32] |
|  |  |  |  |  |  |  |  | BACS | 0.76 | Null/Anecdotal | 0.34 [-0.04, 0.69] |
|  |  |  |  |  |  |  |  | MSCEIT | 1.52 | Alternative/Anecdotal | -0.45 [-0.83, -0.06]\* |
|  |  |  |  |  |  |  |  | Criterion | 0.23 | Null/Moderate | 0.08 [-0.27, 0.43] |
| 5 | 1 | -39.31 | 4.11 | -1.08 | 0.6 | 0.26 | Null/Moderate |  |  |  |  |
|  |  |  |  |  |  |  |  | Intercept | 0.19 | Null/Moderate | 0 [-0.3, 0.33] |
|  |  |  |  |  |  |  |  | BACS | 0.88 | Null/Anecdotal | 0.37 [-0.01, 0.76] |
|  |  |  |  |  |  |  |  | MSCEIT | 1.33 | Alternative/Anecdotal | -0.4 [-0.75, -0.04]\* |
|  |  |  |  |  |  |  |  | Discriminability | 0.25 | Null/Moderate | -0.12 [-0.49, 0.25] |

##### 20.3.5 Traditional Metrics as Predictors (HC Only)

Bayesian Regression: Predicting Social Functioning from Traditional
Metrics (Within HC Only)

| Model | Ref. Model | LOO ELPD | LOO ELPD SE | ΔELPD | ΔELPD SE | Model BF | Model BF Strength/ Direction | Predictor | Pred BF | Pred BF Strength/ Direction | Pred Mean [90% HDI] |
| --- | --- | --- | --- | --- | --- | --- | --- | --- | --- | --- | --- |
| 1 | 1 | -26.22 | 2.11 | – | – | – | –/– |  |  |  |  |
|  |  |  |  |  |  |  |  | Intercept | 0.22 | Null/Moderate | 0 [-0.39, 0.35] |
|  |  |  |  |  |  |  |  | BACS | 0.24 | Null/Moderate | -0.09 [-0.47, 0.3] |
|  |  |  |  |  |  |  |  | MSCEIT | 2.19 | Alternative/Anecdotal | 0.48 [0.1, 0.86]\* |
| 2 | 1 | -25.47 | 2.38 | 0.75 | 1.01 | 1.12 | Alternative/Anecdotal |  |  |  |  |
|  |  |  |  |  |  |  |  | Intercept | 0.2 | Null/Moderate | 0 [-0.35, 0.36] |
|  |  |  |  |  |  |  |  | BACS | 1.02 | Alternative/Anecdotal | -0.53 [-1.16, 0.11] |
|  |  |  |  |  |  |  |  | MSCEIT | 4.32 | Alternative/Moderate | 0.55 [0.16, 0.91]\* |
|  |  |  |  |  |  |  |  | Accuracy | 1.12 | Alternative/Anecdotal | 0.55 [-0.13, 1.16] |
| 3 | 1 | -27.53 | 2.08 | -1.31 | 0.62 | 0.26 | Null/Moderate |  |  |  |  |
|  |  |  |  |  |  |  |  | Intercept | 0.22 | Null/Moderate | 0 [-0.39, 0.38] |
|  |  |  |  |  |  |  |  | BACS | 0.27 | Null/Moderate | -0.11 [-0.51, 0.3] |
|  |  |  |  |  |  |  |  | MSCEIT | 1.76 | Alternative/Anecdotal | 0.47 [0.06, 0.85]\* |
|  |  |  |  |  |  |  |  | RT | 0.27 | Null/Moderate | 0.09 [-0.31, 0.51] |
| 4 | 1 | -26.57 | 2.1 | -0.36 | 0.72 | 0.44 | Null/Anecdotal |  |  |  |  |
|  |  |  |  |  |  |  |  | Intercept | 0.22 | Null/Moderate | 0 [-0.37, 0.36] |
|  |  |  |  |  |  |  |  | BACS | 0.33 | Null/Anecdotal | -0.19 [-0.61, 0.23] |
|  |  |  |  |  |  |  |  | MSCEIT | 1.01 | Alternative/Anecdotal | 0.4 [-0.01, 0.79] |
|  |  |  |  |  |  |  |  | Criterion | 0.43 | Null/Anecdotal | -0.26 [-0.7, 0.16] |
| 5 | 1 | -27.23 | 2.23 | -1.01 | 0.3 | 0.41 | Null/Anecdotal |  |  |  |  |
|  |  |  |  |  |  |  |  | Intercept | 0.22 | Null/Moderate | 0 [-0.39, 0.37] |
|  |  |  |  |  |  |  |  | BACS | 0.42 | Null/Anecdotal | -0.06 [-0.76, 0.63] |
|  |  |  |  |  |  |  |  | MSCEIT | 1.78 | Alternative/Anecdotal | 0.47 [0.06, 0.86]\* |
|  |  |  |  |  |  |  |  | Discriminability | 0.42 | Null/Anecdotal | -0.03 [-0.7, 0.68] |
